# Large-scale evaluation of proteomic and polygenic risk scores reveals complementary contributions to incident disease prediction

**DOI:** 10.1101/2025.07.10.25331242

**Authors:** Jakob Woerner, Thomas M. Westbrook, Jaehyun Joo, Manu Shivakumar, Rasika Venkatesh, Tess Cherlin, Sang-Hyuk Jung, Seokho Jeong, Damian Maseda, Michelle McKeague, Shwetank, Matei Ionita, Joost Wagenaar, Sarah A. Abramowitz, Anurag Verma, Bingxin Zhao, Seunggeun Lee, Scott Damrauer, Michael G. Levin, Su Chin Heo, Thomas P. Cappola, Daniel J. Rader, Sharlene Day, Rajat Deo, Joel M. Gelfand, Ravi Ramessur, Marie A. Guerraty, Shefali Setia-Verma, Bogdan Pasaniuc, Marylyn D. Ritchie, Sokratis A. Apostolidis, Allison R. Greenplate, E. John Wherry, Penn Medicine Biobank, Yonghyun Nam, Dokyoon Kim

**Affiliations:** Department of Biostatistics, Epidemiology and Informatics, Perelman School of Medicine, University of Pennsylvania, Philadelphia, PA, 19104, USA; Department of Genetics, University of Pennsylvania Perelman School of Medicine, Philadelphia, Pennsylvania, USA; Department of Pathology and Laboratory Medicine, University of Pennsylvania Perelman School of Medicine, Philadelphia, Pennsylvania, USA; Department of Medical Informatics, Kangwon National University College of Medicine, Chuncheon 24341, Republic of Korea; Graduate School of Data Science, Seoul National University, Seoul, 08826, Republic of Korea; Institute for Immunology & Immune Health (I3H), Perelman School of Medicine, University of Pennsylvania, Philadelphia, PA 19104, USA; Department of Surgery, University of Pennsylvania Perelman School of Medicine, Philadelphia, PA, 19104, USA; Division of Translational Medicine and Human Genetics, Department of Medicine, University of Pennsylvania - Perelman School of Medicine, Philadelphia, PA 19104, USA; Institute for Biomedical Informatics, University of Pennsylvania - Perelman School of Medicine, Philadelphia, PA 19104, USA; Department of Statistics and Data Science, University of Pennsylvania, Philadelphia, PA 19104, USA; Cardiovascular Institute, University of Pennsylvania Perelman School of Medicine, Philadelphia, PA, 19104, USA; Department of Surgery, Corporal Michael J. Crescenz VA Medical Center, Philadelphia, PA, 19104, USA; Division of Cardiovascular Medicine, Department of Medicine, University of Pennsylvania Perelman School of Medicine, Philadelphia, PA, 19104, USA; Department of Medicine, Corporal Michael J. Crescenz VA Medical Center, Philadelphia, PA, 19104, USA; McKay Orthopaedic Research Laboratory, Department of Orthopaedic Surgery, Perelman School of Medicine, University of Pennsylvania, Philadelphia, PA, 19104, USA; Translational Musculoskeletal Research Center, Corporal Michael J Crescenz VA Medical Center, Philadelphia, PA, 19104, USA; Division of Cardiovascular Medicine, Hospital of The University of Pennsylvania, Philadelphia, Pennsylvania; Division of Cardiovascular Medicine, Electrophysiology Section, Hospital of the University of Pennsylvania, Philadelphia, Pennsylvania, USA; St John’s Institute of Dermatology, School of Basic & Medical Biosciences, Faculty of Life Sciences & Medicine, King’s College London, London, United Kingdom; Department of Medicine, University of Pennsylvania Perelman School of Medicine, 3400 Civic Center Blvd., Philadelphia, PA 19104, USA; Division of Rheumatology, Department of Medicine, Perelman School of Medicine, University of Pennsylvania, Philadelphia, PA 19104, USA; Department of Systems Pharmacology and Translational Therapeutics, Perelman School of Medicine, University of Pennsylvania, Philadelphia, PA 19104, USA; Parker Institute for Cancer Immunotherapy, Perelman School of Medicine, University of Pennsylvania, Philadelphia, PA 19104, USA

## Abstract

Plasma proteins capture dynamic physiological processes and may offer more immediate insight into disease risk than static genetic predictors. We evaluated the predictive utility of proteomic risk scores (ProRS) versus polygenic risk scores (PRS) across 301 phenotypes in 39,843 participants from the UK Biobank Pharma Proteomics Project. ProRS, trained on prevalent cases, were tested for incident disease and benchmarked against PRS derived from genome-wide association statistics. Among 268 phenotypes with informative signals, ProRS outperformed PRS in 88% of traits (a median C-index improvement of 9.6%), showing strongest gains for circulatory, metabolic, and immune conditions. Combined models further improved prediction, particularly for traits with higher heritability. Longitudinal analyses showed that ProRS values were elevated years before diagnosis. External validation in 841 Penn Medicine BioBank participants confirmed consistent performance and transferability, with AUC improvements up to 4.18% over PRS alone. Plasma proteomic profiling provides complementary, temporally responsive information that enhances individual-level disease prediction.

## Introduction

Large-scale plasma proteomics is redefining our understanding of human disease biology and enabling new avenues for risk prediction. Landmark efforts, including the UK Biobank (UKB) proteomics atlas profiling 2,920 proteins across >53,000 individuals, have uncovered widespread associations between circulating proteins and hundreds of prevalent and incident diseases, reinforcing the plasma proteome’s role as a dynamic indicator of health status^1–3^. Recent proteomics studies have demonstrated the clinical relevance of proteomic aging clocks^4^, predictive signatures for common and rare diseases^5^, and integrated models of aging^6–8^. Proteomics has also been applied to predict impaired glucose tolerance^9^, cardiorespiratory fitness^10^, and β-amyloid pathology after SARS-CoV-2 infection^11^, further underscoring its value for early disease detection.

Despite this progress, most translational applications have focused on individual outcomes or trait-specific protein signatures^12–14^. Polygenic risk scores (PRS) offer fixed, inherited estimates of genetic predisposition and are now widely used across biobank studies. However, they cannot capture dynamic physiological changes that may precede clinical disease. In contrast, proteomic risk scores (ProRS), derived from plasma protein levels, are sensitive to time-varying physiological changes, reflect downstream effects of genetic traits, and may capture subclinical molecular perturbations that provide greater predictive power before the clinical onset of disease. While PRS and ProRS are inherently complementary, no prior studies have systematically evaluated their additive and interactive contributions to incident disease prediction across the phenome^15^.

Here, we address this need by jointly modeling PRS and ProRS across 301 disease phenotypes in 39,843 participants from the UK Biobank Pharma Proteomics Project (UKB-PPP), enabling a systematic evaluation of their predictive performance across a wide range of clinical conditions. ProRS models were developed using prevalent cases and evaluated for incident disease prediction, capturing a clinically relevant trajectory from health to diagnosis. We quantified the independent and combined predictive value of PRS and ProRS across the phenome, analyzed the temporal dynamics of proteomic risk, examined cross-trait relationships between proteomic and genetic risk scores, and validated model generalizability in an external cohort. Our findings establish a scalable framework for integrated risk stratification and provide a foundation for advanced precision prediction using combined high-dimensional proteomic and genomic data across multiple disease states.

## Results

### Phenome-wide development of PRS and ProRS across 301 binary phenotypes

We analyzed data from 39,843 unrelated individuals with high genetic similarity with European reference populations from UKB-PPP, with baseline plasma proteomics measured using the Olink Explore 3072 panel and genome-wide genotype data imputed using the UKB pipeline (**Fig. 1**). The median follow-up time was 13.8 years (**Supplementary Table 1**). Disease phenotypes were defined by integrating ICD-coded diagnoses from inpatient and primary care records, cancer registries, and death records, using endpoint definitions adapted from the FinnGen project. Participants were classified as prevalent cases (diagnosed on or before baseline), incident cases (diagnosed after baseline), or controls (no diagnosis as of November 2023) (**Fig. 1a**). Sensitivity analyses were conducted to ensure accurate identification of truly incident cases. Starting from 2,466 phenotypes with available GWAS summary statistics from FinnGen, we included 301 binary phenotypes with at least 30 prevalent and 30 incident cases for downstream modeling (**Supplementary Fig. 1, Supplementary Table 2**). For each phenotype, the cohort was partitioned into individuals with prevalent disease at baseline and randomly sampled controls for model training, and those with incident disease and the remaining controls for evaluation.

**Fig. 1.**
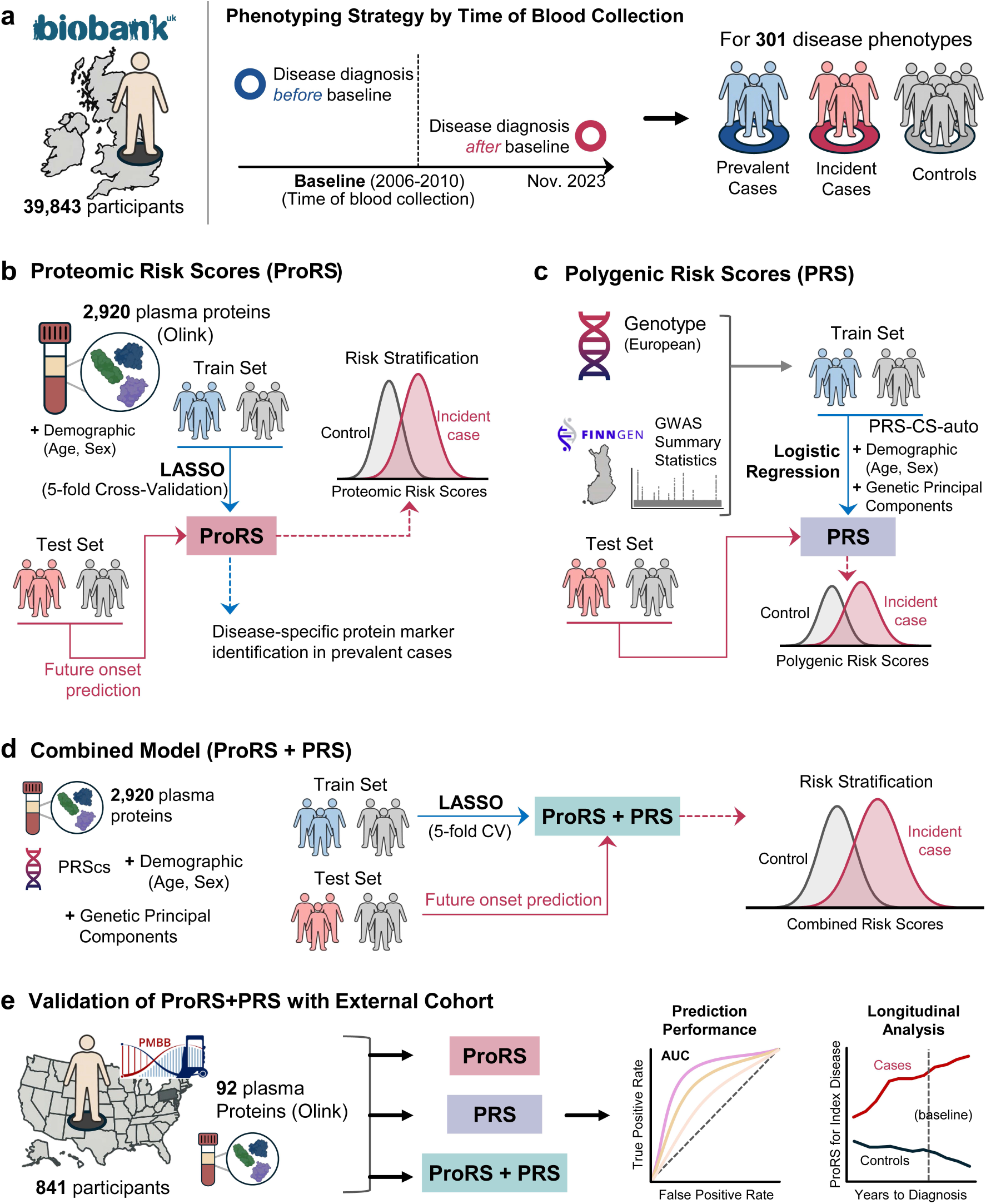
Study design and modeling framework for incident disease prediction using proteomic and genetic risk scores. **a**, Overview of cohort and phenotyping strategy. We analyzed 39,843 participants from the UK Biobank Pharma Proteomics Project (UKB-PPP) with baseline plasma proteomics (2006–2010) and median follow-up of 13.8 years. For each of 301 disease phenotypes, individuals were classified as prevalent cases (diagnosed before baseline), incident cases (diagnosed after baseline), or controls. **b**, Proteomic risk scores (ProRS) were trained using 2,920 plasma proteins from prevalent cases and controls, with five-fold cross- validation, and tested in incident cases. **c**, Polygenic risk scores (PRS) were computed using FinnGen GWAS summary statistics and adjusted for demographic and genetic covariates in logistic regression models. **d**, Combined models (PRS+ ProRS) were trained using LASSO with integrated proteomic, genetic, and demographic features. **e**, External validation was conducted in 841 participants from the Penn Medicine Biobank using 92 overlapping proteins, evaluating prediction performance and longitudinal trajectories of ProRS.

ProRS were generated using 2,920 plasma proteins passing quality control (**Methods**). For each phenotype, L1-regularized (LASSO) logistic regression models were trained to distinguish prevalent cases from controls, adjusting for age and sex^16^. Five-fold cross-validation was used for model tuning and feature selection (**Fig. 1b**). PRS were constructed using summary statistics from the FinnGen study (release 12)^17^, with single-nucleotide polymorphisms (SNP) weights derived using PRS-CS-auto, a Bayesian method that applies continuous shrinkage priors and automatically tunes the global shrinkage parameter, restricted to ∼1.2 million HapMap3 variants^18^. PRS were computed for each individual and adjusted by age, sex, and the top 10 genetic principal components with logistic regression (**Fig. 1c**). Combined models (PRS+ProRS) were developed using LASSO logistic regression with PRS and 2,920 plasma proteins with age, sex, and genetic PCs as covariates (**Fig. 1d**)^19^. All models were trained on prevalent cases and tested on independent sets of incident cases and controls (**Supplementary Table 3**). This approach allowed systematic evaluation of static genetic and dynamic proteomic risk layers in the context of incident disease prediction.

To assess generalizability, we conducted external validation in 841 individuals from the Penn Medicine BioBank (PMBB)^20^ with both genotype data and proteomic measurements from the Olink Target 96 Cardiovascular II panel. For this, ProRS models were retrained in UKB using only the 92 proteins available in PMBB and applied without recalibration (**Fig. 1e**).

### Proteomic scores outperform genetic scores across the phenome

We systematically benchmarked the predictive performance of ProRS against PRS across 301 diseases, evaluating all models using Harrell’s concordance index (C-index) to quantify discrimination in incident-only prediction settings^21^. Among these, 268 phenotypes had at least one protein selected by the ProRS or combined (PRS+ProRS) model and were included in comparative analyses. ProRS yielded higher predictive accuracy than PRS in 88.8% of these traits (median C-index improvement = 9.6%; IQR = 3.8%- 14.9%; **Fig. 2a, Supplementary Table 4**), a modest but consistent gain that may offer meaningful utility in stratifying population-level risk across diverse disease domains. C-index values from ProRS and PRS were moderately aligned across the phenome (R^2^ = 0.5417) while individual-level correlations between the two scores were generally low across phenotypes (**Supplementary Fig. 2**), suggesting that ProRS and PRS capture complementary aspects of disease risk.

**Fig. 2.**
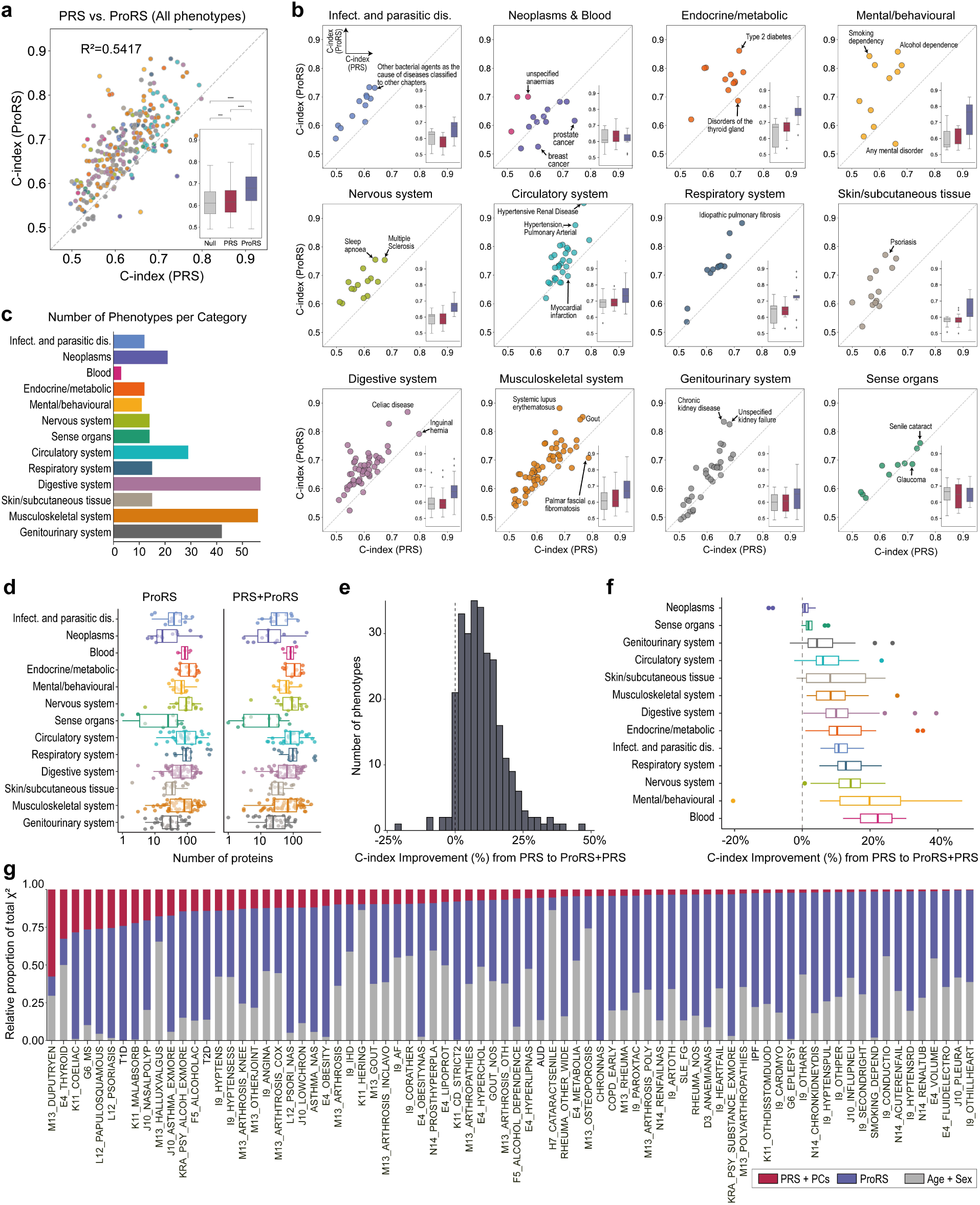
Comparative performance of proteomic and genetic risk scores across the phenome. **a,** C-index comparison between ProRS and PRS models across 268 phenotypes with at least one protein selected by either model. Inset: C-index distributions for null (age + sex), PRS, and ProRS models. **b,** ProRS vs. PRS performance stratified by 13 clinical phenotype categories; selected traits labeled. Inset: category-specific C-index box plots. **c,** Number of phenotypes per category among the 268 included in the analysis. **d,** Number of proteins selected in ProRS-only and combined models, stratified by phenotype category. Non-zero LASSO coefficients; see **Supplementary Tables 7 and 8**. **e,** Histogram of relative improvement in C-index from PRS to combined model. **f**, Category-stratified box plots of C-index improvement; modest declines observed in neoplastic and immune-related traits. **g,** χ² decomposition of variance explained (Nagelkerke R² > 0.1), showing relative contributions of ProRS, PRS (including PCs), and age/sex covariates. ProRS explained the largest proportion of variance in most traits; PRS contributed more in autoimmune-related phenotypes.

When stratified by clinical phenotype category, ProRS consistently outperformed PRS across most of the 13 disease systems evaluated (**Fig. 2b**). Gains were particularly evident for diseases of the circulatory, respiratory, digestive, and skin/subcutaneous systems. For example, idiopathic pulmonary fibrosis, myocardial infarction, celiac disease, and psoriasis exhibited high overall predictability, with ProRS consistently outperforming PRS. While these diseases are influenced by both genetic and non-genetic factors, the performance gains from ProRS suggest added value in capturing dynamic, preclinical risk signals beyond static genetic predisposition. In contrast, PRS showed stronger performance in most neoplastic phenotypes, including breast and prostate cancer, where genetic risk is known to play a more dominant role. For breast and prostate cancer, PRS outperformed ProRS by 8.5% (C-index: 0.6106 vs. 0.5256) and 12.6% (0.7423 vs. 0.6166), respectively. Notably, the combined (PRS+ProRS) model did not improve prediction for these two cancers, which appear to be exceptions to the broader phenome-wide trend. The number of phenotypes per category is summarized in **Fig. 2c**.

We next examined the added value of ProRS when integrated with PRS. Across 268 phenotypes, the number of proteins selected by the ProRS-only and combined (PRS+ProRS) models was broadly similar, with modest reductions in the number of selected proteins observed in some categories after inclusion of genetic predictors (**Fig. 2d, Supplementary Table 5**). The addition of ProRS consistently enhanced predictive performance beyond PRS alone, reinforcing the complementary predictive value of proteomic and genetic risk scores. Across the phenome, 94.8% of traits showed improved predictive performance with the addition of ProRS, with C-index improvements up to 47.2% (median=8.6%, IQR=4.1%-13.8%). While most phenotypes benefited from the combined model (**Fig. 2e**), a minority of traits (14 out of 268; 5.2%) showed minimal or no gain, with slight decreases observed in some neoplastic (e.g., breast and prostate cancer), circulatory (e.g., myocardial infarction, stroke), and genitourinary phenotypes (e.g., ovarian cyst, postmenopausal bleeding) (**Fig. 2f**). To better understand the contribution of each component, we decomposed model variance (Nagelkerke R² > 0.1) using χ² analysis (**Fig. 2g**). Using a Cox proportional hazards model with PRS, ProRS, and covariates as predictors, we partitioned the total model variance into χ² components via analysis of variance^22^. Across most phenotypes, ProRS accounted for the largest share of explained variance, underscoring its dominant role in individual-level prediction. Notably, for the eight phenotypes in which the PRS explained the largest proportion of model variance (adjusted by ancestry via genetic PCs), all were autoimmune diseases (e.g., type 1 diabetes, multiple sclerosis, celiac disease, and psoriasis) or related comorbidities such as intestinal malabsorption and thyroid disorders (**Supplementary Table 6**).

Despite the overall superiority of ProRS in predicting incident disease, the added benefit of incorporating PRS varied across traits. To examine whether this variability reflects underlying genetic architecture, we assessed SNP-based heritability (h²) using LD score regression (LDSC) from the FinnGen GWAS data used to construct PRSs^23^. We found that differences in protein selection and performance gain between ProRS and combined models were associated with h² (**Extended Data Fig. 1**). Among the 268 phenotypes analyzed, 87% showed a change in protein selection with PRS inclusion, and 53% had fewer proteins selected by the combined model (**Supplementary Fig. 3**), with reductions most pronounced in traits with higher heritability (**Extended Data Fig. 1a**), suggesting that the predictive contribution of PRS may overlap with proteomic features, particularly in traits with stronger genetic architecture. In contrast, protein selection remained stable in traits with low heritability (h² < 0.02), where PRS offered limited predictive value. C-index improvements from ProRS to PRS+ProRS were positively correlated with heritability (Spearman ρ = 0.56; **Extended Data Fig. 1b**), with minimal or negative gains observed for low- h² traits. Stratifying phenotypes by heritability tiers confirmed these trends (**Extended Data Fig. 1c)**. In traits with h² < 0.02, PRS performed similarly to the covariate-only model, and the combined model added little beyond ProRS. In contrast, for h² > 0.04, PRS improved prediction and enhanced the combined model’s performance. Category-level analysis revealed that PRS was most beneficial in more heritable traits such as neoplasms, while ProRS maintained stable performance across all categories, including those with lower heritability (**Supplementary Fig. 4**). These findings demonstrate that PRS adds value primarily in traits with stronger genetic contribution, whereas ProRS provides robust prediction across a broader range of diseases.

### Complementary risk stratification from genetic and proteomic scores in complex diseases

To evaluate the translational potential of integrating genomic and proteomic risk models, we assessed whether combining PRS and ProRS improves individual-level stratification across disease categories. We focused on twelve representative traits based on sample size, clinical relevance, and diversity of underlying pathophysiology. This panel included six Cardiovascular-Kidney-Metabolic (CKM) traits^24^— AF, coronary heart disease (CHD), myocardial infarction (MI), venous thromboembolism (VTE), CKD, and type 2 diabetes (T2D)—and six Autoimmune Diseases (AiD): multiple sclerosis (MS), celiac disease (CED), ulcerative colitis (UC), psoriasis (PSO), rheumatoid arthritis (RA), and systemic lupus erythematosus (SLE) (**Supplementary Table 9**). These traits span a range of genetic and epidemiologic profiles, including variation in case counts, disease prevalence, and underlying biological pathways^25–27^.

Across twelve diseases, the combined (PRS+ProRS) model achieved the highest predictive performance in most cases. It improved AUC in 8 of the 12 traits with an average increase of 1.8% (range: 0.18 to 3.17%) over the stronger of the two single-modality models (**Figs. 3a-3b**; **Supplementary Table 10**). In the remaining four traits (CHD, MI, RA, and SLE), predictive performance was comparable, with only minor decreases (<1.1%). ProRS outperformed PRS in most CKM traits, with notable advantages in AF, CKD, VTE, and T2D. In contrast, PRS slightly outperformed ProRS in CHD and MI. For all AiDs, ProRS consistently achieved higher predictive accuracy than PRS.

**Fig. 3.**
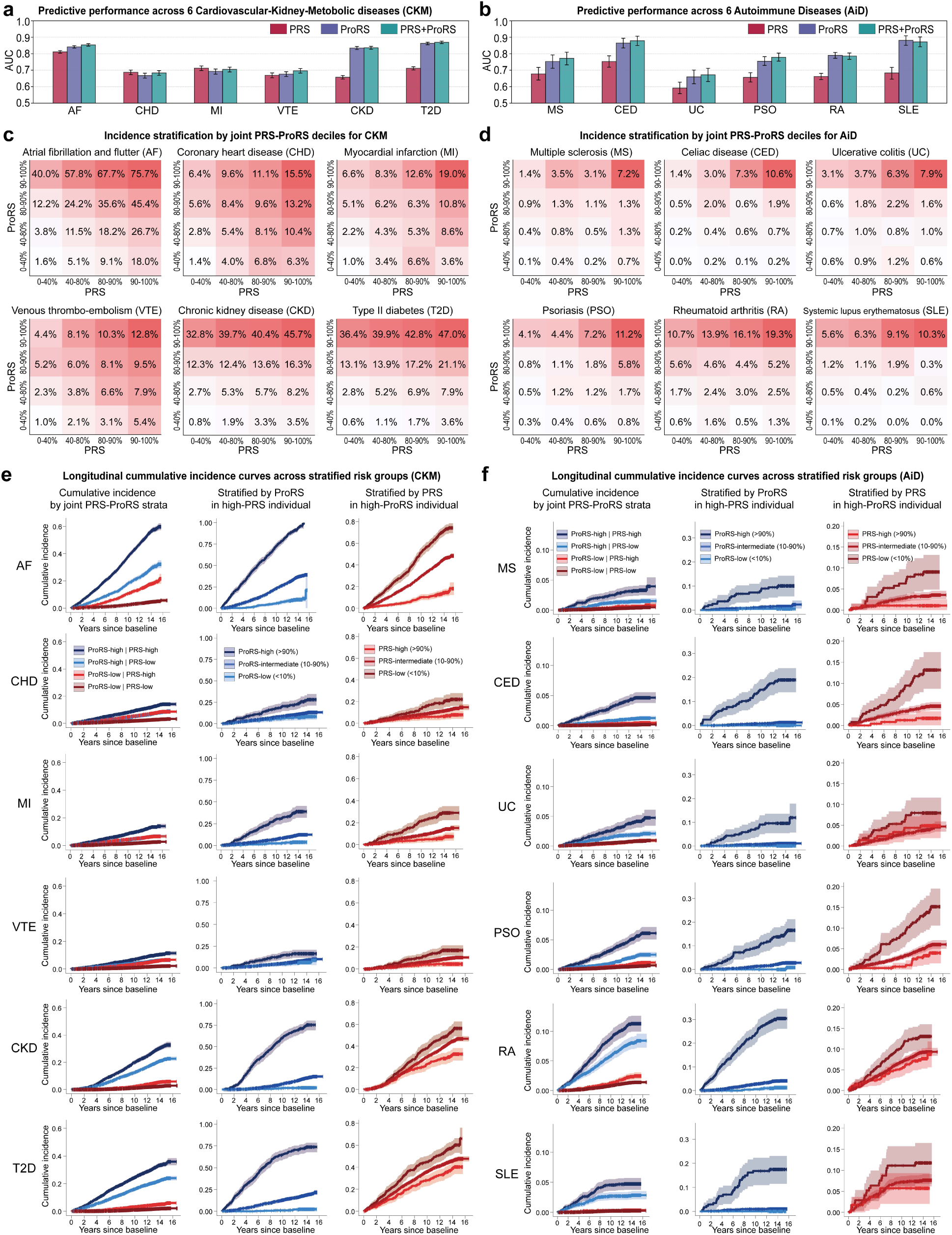
Complementary contributions of PRS and ProRS to disease prediction and risk stratification. **a–b**, Predictive performance across six Cardiovascular-Kidney-Metabolic (CKM; a) and six autoimmune diseases (AiDs; b), showing average AUC for PRS, ProRS, and combined (PRS+ProRS). Error bars represent 95% confidence intervals estimated by bootstrap resampling; full estimates are reported in **Supplementary Table 10. c–d**, Stratified incidence heatmaps by joint PRS–ProRS deciles for CKM (c) and AiD (d) traits. The x-axis and y-axis represent four percentile strata (0–40%, 40–80%, 80–90%, 90–100%) of PRS and ProRS, respectively. Each cell indicates the proportion of individuals diagnosed with the disease during the follow-up period, with incidence rates rising progressively along both PRS and ProRS axes. **e–f**, Cumulative incidence curves across stratified subgroups for CKM (e) and AiD (f) traits. Left panels show incidence across four joint PRS–ProRS strata (high = top 25%); corresponding log-rank test results are reported in **Supplementary Table 11**. Middle panels show ProRS-based stratification within individuals in the top 25% of PRS; log-rank results in **Supplementary Table 12**. Right panels show PRS-based stratification within individuals in the top 25% of ProRS; log-rank results in **Supplementary Table 13**.

To evaluate the stratification utility of combining PRS and ProRS at the individual level, we partitioned participants into 16 subgroups based on joint percentiles of both scores: 0–40%, 40–80%, 80– 90%, and 90–100% along each axis. Cumulative disease incidence was then estimated within each subgroup (**Figs. 3c-3d**), capturing a risk gradient from low to high. Across all twelve traits, incidence increased along both axes, with the highest values observed in the 90–100th percentile for both PRS and ProRS. In this high-risk stratum, cumulative incidence reached 75.7% for AF and 47.0% for T2D. A stepwise gradient in incidence was observed as both scores increased, particularly in CKD, AF, and T2D. Notably, elevated risk was also present in subgroups where only one of the two scores was high. For example, in AF, individuals with high ProRS (90–100th percentile) but low PRS (0–40th percentile) had a cumulative incidence of 40.0%, whereas those with high PRS and low ProRS had an incidence of 18.0%. These patterns suggest that each score contributes independent and complementary risk information. Similar trends were observed in other diseases, though the distribution of incidence varied by trait. While incidence was generally lower for AiDs than CKM traits due to their lower prevalence, stratified patterns remained evident, supporting the utility of combined risk models across disease domains.

To examine how PRS and ProRS jointly influence disease risk over time, we evaluated cumulative incidence curves across stratified subgroups for each CKM (**Fig. 3e**) and AiD trait (**Fig. 3f**). In the first analysis (left panels), individuals were classified into four groups based on binary thresholds for each score (high >75th percentile, low ≤75th percentile) to assess joint risk effects. Across both disease domains, individuals with high PRS and high ProRS exhibited the steepest cumulative incidence curves, while those with lower scores on one or both axes had consistently attenuated risk profiles. This divergence was particularly strong among CKM traits where cumulative incidence curves began to separate early and widened over time (**Fig. 3e, left**). For AiDs, stratified curves also revealed consistent separation across risk groups, with individuals in the joint high-risk group (high PRS and high ProRS risk group) showing the highest cumulative incidence, particularly in conditions such as CED and RA. These patterns highlight the additive value of combining genetic and proteomic scores (**Fig. 3f, left**).

To further disentangle the contribution of each modality, we examined incidence within high-risk subsets for one score while varying the other. Among individuals with high PRS (>75th percentile), stratification by ProRS revealed additional risk gradation in several traits (**Figs. 3e** and **3f, middle**), with particularly strong separation observed in RA. A complementary pattern emerged when stratifying PRS within those with high ProRS: in traits such as AF, MS, and SLE, incidence curves showed clear separation between PRS strata, even among individuals with similarly elevated ProRS (**Figs. 3e** and **3f, right**). ProRS can capture temporally proximal or active disease processes across multiple traits, while PRS enhances resolution when integrated, particularly in settings with sufficient case numbers to support subgrouping.

### Proteomic risk scores capture dynamic trajectories preceding disease onset

A fundamental distinction between PRS and ProRS lies in their sensitivity to changes over time. While the underlying genetic liability captured by PRS is fixed from birth, ProRS reflects the evolving physiological state of an individual. Protein levels can capture a combination of genetic and non-genetic influences, but unlike PRS, they are sensitive to modifiable factors such as environment, lifestyle, and therapeutic interventions^28^. To examine this temporal dimension, we assessed the time-dependent predictive performance and longitudinal dynamics of ProRS relative to disease onset across 12 traits spanning cardiometabolic, renal, and autoimmune domains (**Fig. 4**).

**Fig. 4.**
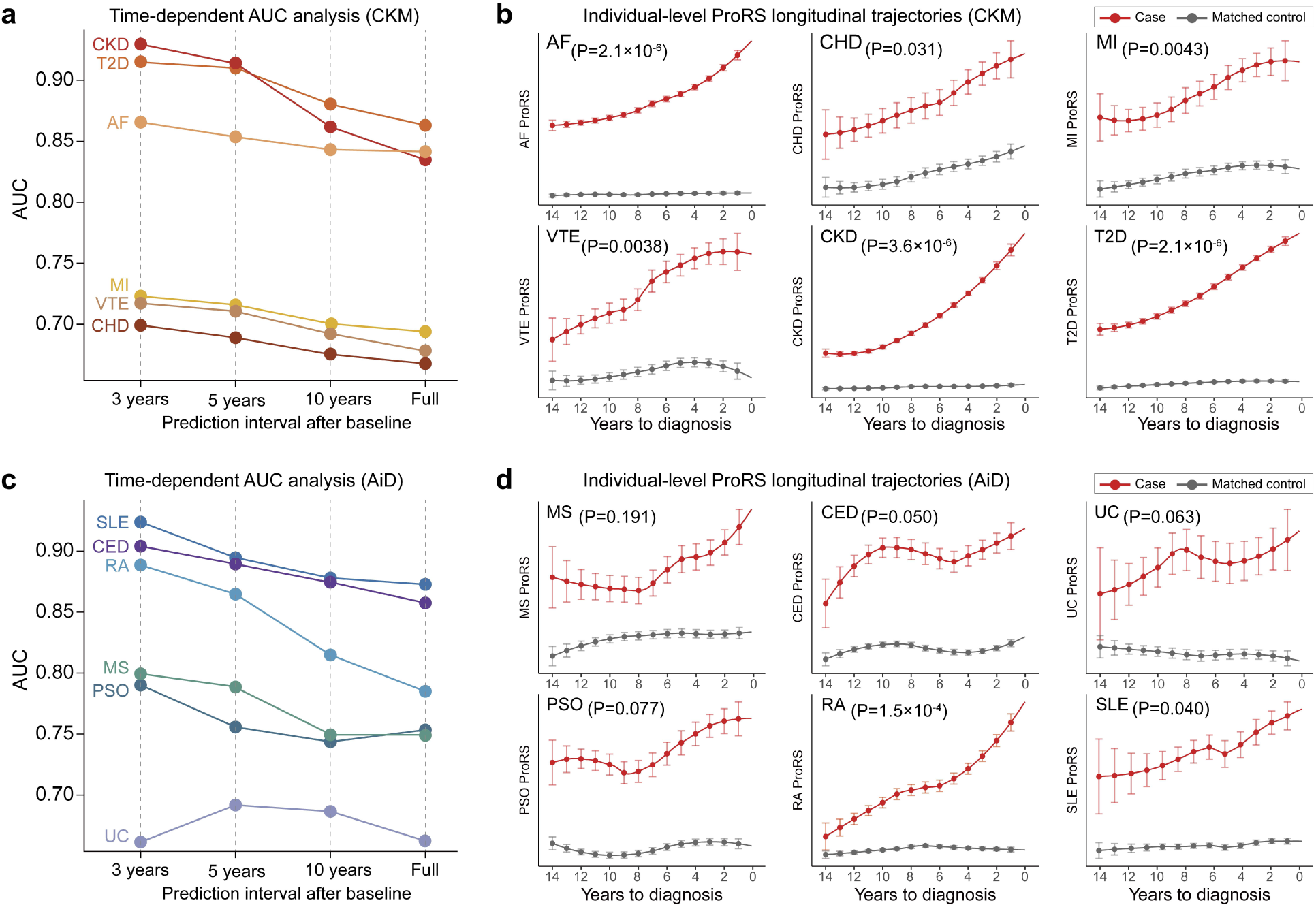
Temporal performance and preclinical trajectories of ProRS. **a, c**, Time-dependent AUCs for ProRS across six Cardiovascular-Kidney-Metabolic traits (CKM; a) and six autoimmune diseases (AiDs; c), evaluated at 3, 5, and 10 years after baseline and over the full follow-up (median=13.8 years). ProRS performance declined with increasing duration from sampling and analysis of blood, highlighting stronger predictive value closer to diagnosis. **b, d**, Longitudinal ProRS trajectories in cases (red) against age- and sex-matched controls (gray) for CKM (b) and AiD traits (d), aligned to years prior to diagnosis. ProRS values increased steadily as diagnosis approached in most traits, with elevated levels often evident over a decade in advance. P-values are from the Mann-Kendall trend test on case–control differences.

We first assessed predictive performance across four time windows: 3, 5, and 10 years after baseline, as well as the full follow-up (median = 13.8 years). Across nearly all 12 traits, ProRS achieved the highest AUCs at shorter time horizons, with performance gradually declining over longer intervals (**Figs. 4a** and **4c**). For example, AUCs for CKD, T2D, and RA exceeded 0.90 within 3 years of baseline, but decreased when predictions were extended across the full follow-up period. These results demonstrate that ProRS more accurately predicts disease risk in shorter-term intervals, consistent with temporally proximate changes in circulating protein levels.

To evaluate how ProRS evolves in the years preceding clinical onset, we analyzed its longitudinal trajectories in cases versus age- and sex-matched controls across single-year time intervals prior to diagnosis (**Figs. 4b** and **4d**). Incident cases were grouped by duration from baseline sampling to clinical diagnosis, and controls were anchored to the same timeline using a nested case-control framework. Across all CKM and AiD traits, ProRS levels rose progressively as diagnosis approached. Importantly, elevated scores were often detectable more than a decade before clinical onset, underscoring the temporal sensitivity of proteomic profiles. This pattern was especially pronounced in CKD, T2D, and RA, which exhibited monotonic increases in ProRS years prior to diagnosis Individuals diagnosed with CKD as late as 14 years after baseline already showed significantly elevated ProRS compared to age- and sex matched controls, and ProRS levels increased steadily with proximity to diagnosis (P=3.6×10⁻⁶, Mann-Kendall trend test; **Fig. 4b**). ProRS trajectories increased consistently prior to diagnosis, particularly in conditions with slow or subclinical progression such as CKD, T2D, and RA, where early pathophysiological changes precede clinical recognition. While prediction performance generally declined with longer horizons, ProRS retained meaningful discriminative power even a decade before diagnosis, supporting its potential utility across both chronic and more acute disease contexts.

### Genetic–proteomic risk score interaction does not drive prediction performance

To assess whether combined predictive effects of PRS and ProRS extend beyond additivity, we included a multiplicative interaction term in the Cox model. Among 268 traits, 21 showed significant interactions after FDR correction (FDR < 0.05; **Supplementary Table 14**), with the strongest effects in hypertension (FDR = 4.4 × 10⁻³⁰) and T2D. In these cases, one modality modulated the impact of the other (**Supplementary Fig. 5)**. However, most traits exhibited minimal interaction, and additive models generally sufficed. While synergistic effects may exist for select conditions, additive integration remains appropriate for most phenotypes.

### Associations between ProRS and PRS reveal shared cross-trait risk patterns

While PRS-based cross-trait associations are well-studied^29^, the added dimension of proteomic profiles may offer complementary insight into shared biological pathways and comorbid risk across clinical domains. We conducted a phenome-wide association analysis between 246 ProRSs and corresponding PRSs to assess where proteomic and genetic risk converge or diverge (**Methods**). As expected, the strongest associations were typically observed for the same trait, where each ProRS aligned with its corresponding PRS. We also identified numerous significant cross-trait associations, in which the ProRS for one disease was associated with the PRS of a different condition, including links between phenotypes from disparate clinical categories. The P-values and corresponding effect sizes for each PRS–ProRS pair are reported in **Supplementary Table 15**.

To illustrate these patterns, we examined three representative traits: obesity, AF, and CKD. The obesity PRS was strongly associated with ProRSs for cardiometabolic and inflammatory traits including sleep apnea, type 2 diabetes, cellulitis, and arthrosis (**Fig. 5a**). The AF PRS correlated with ProRSs for heart failure and pulmonary conditions such as COPD and respiratory infections (**Fig. 5b)**, while the CKD PRS aligned with ProRSs for acute renal failure, anemia, and sepsis (**Fig. 5c**). These cross-domain links suggest that ProRS captures downstream physiological pathways that are partly shaped by genetic risk but also reflect broader comorbid processes.

**Fig. 5.**
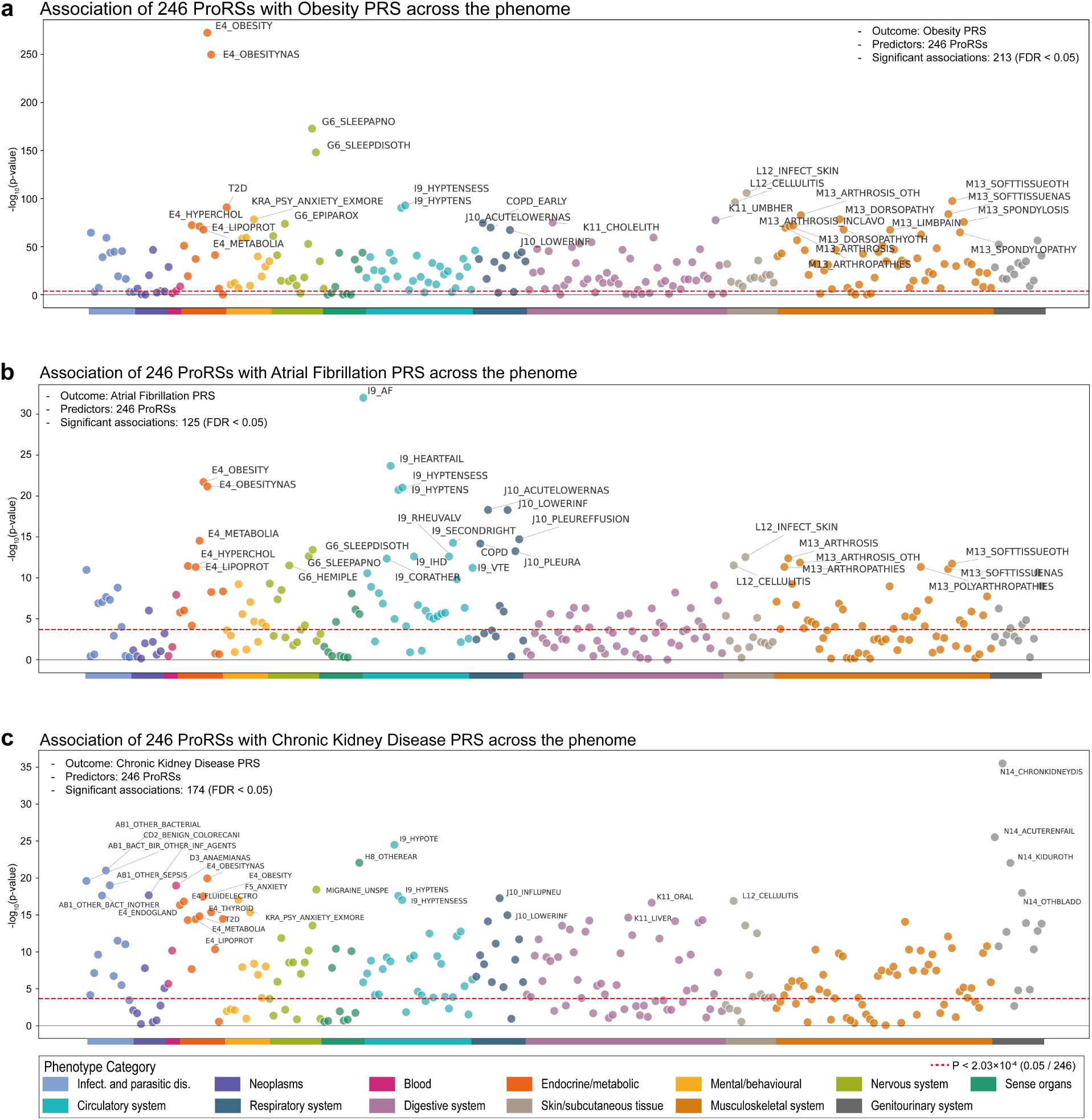
Cross-trait associations between proteomic and genetic risk scores for selected traits. **a–c**, For three selected traits, we modeled the polygenic risk score (PRS) as a function of 246 proteomic risk scores (ProRSs), adjusting for age, sex, and ancestry principal components. Each panel displays –log₁₀(FDR-adjusted P-values) for ProRS predictors, grouped by 13 clinical phenotype categories. Dashed red lines indicate the significance threshold (FDR < 0.05). These examples illustrate the diversity of cross-trait associations observed across the phenome.

### Sensitivity analysis confirms robustness of proteomic predictions at follow-up visits

To assess temporal stability, we retrained ProRS models using protein measurements from UK Biobank follow-up visits (instances 2 and 3), where a reduced set of 1,450 proteins was available. Despite smaller sample sizes and lower protein coverage, predictive performance remained consistent: ProRS outperformed PRS, and combined models showed the highest accuracy across both follow-up points (**Supplementary Figs. 6–7**). Dimensionality reduction confirmed minimal batch effects across visits, supporting reproducibility of proteomic profiles (**Supplementary Fig. 6b**). Population-level protein structure was preserved, though individual trajectories varied, reflecting the dynamic nature of circulating proteomes (**Supplementary Fig. 8**).

We further compared models trained on prevalent versus incident cases, observing high concordance in protein weights (median Spearman ρ = 0.71) and minimal differences in C-index (mean < 0.5%) (**Supplementary Figs. 9–10**). These findings support the generalizability of ProRS models trained on prevalent cases, highlighting the value of future studies incorporating repeated measurements and disease trajectories for refining risk estimation.

### External validation supports generalizability of proteomic and polygenic risk scores

To assess generalizability, we applied our risk prediction models to 841 individuals from the PMBB^20^, a subset of a larger cohort (N ≈ 57,000) with available genotype and plasma proteomic data. Proteins were measured using the Olink Target 96 Cardiovascular II panel, which includes 92 analytes overlapping with those in the UKB-PPP (**Supplementary Table 16**). For harmonized evaluation, we retrained ProRS models in the UKB using only the 92 shared proteins and applied them to PMBB without recalibration. Despite the reduced feature set, predictive performance remained robust, with modest attenuation relative to full-panel models. Both ProRS and combined (PRS+ProRS) models maintained similar predictive accuracy trends across all 268 traits (**Supplementary Table 17**). In UKB, the combined model outperformed ProRS alone in most traits, supporting the rationale for external validation using the 92-protein model and demonstrating its transferability across cohorts.

We first assessed predictive performance across 12 clinical categories in PMBB using AUC (**Fig. 6a; Supplementary Table 18**). ProRS consistently outperformed PRS across multiple systems, and the combined PRS+ProRS model achieved the highest accuracy in most categories. Gains were particularly pronounced in the circulatory, genitourinary, and endocrine/metabolic domains, in concordance with our previous results, where short-term physiological changes may be more effectively captured by circulating protein levels. These findings support the broad applicability of proteomic risk models in external populations and highlight the added predictive value of combining proteomic and genetic risk. To further examine trait-level performance, we focused on CKD, a representative renal condition of clinical relevance for genetic and proteomic risk modeling ^30–32^. In PMBB, the combined model achieved an AUC of 0.84, outperforming ProRS alone (AUC = 0.78), PRS alone (AUC = 0.55), and a covariate-only model (AUC = 0.56) (**Fig. 6b**). Longitudinal analysis of ProRS trajectories revealed a steady increase in proteomic risk in cases during the years preceding diagnosis, while matched controls showed no such trend (P = 0.0098; **Fig. 6c**), consistent with patterns observed in UKB. Decision curve analysis further demonstrated that both ProRS and the combined model yielded higher net benefit than PRS across a range of risk thresholds (**Fig. 6d**)^33,34^, including when benchmarked against estimated glomerular filtration rate (eGFR < 60). Calibration analysis showed close alignment between predicted and observed 5-year risk for ProRS and the combined model, with lower integrated calibration index (ICI) values compared to PRS (**Fig. 6e**). These results underscore the feasibility of applying targeted proteomic panels for population-level risk prediction and support the translational potential of integrated molecular models in diverse clinical cohorts.

**Fig. 6.**
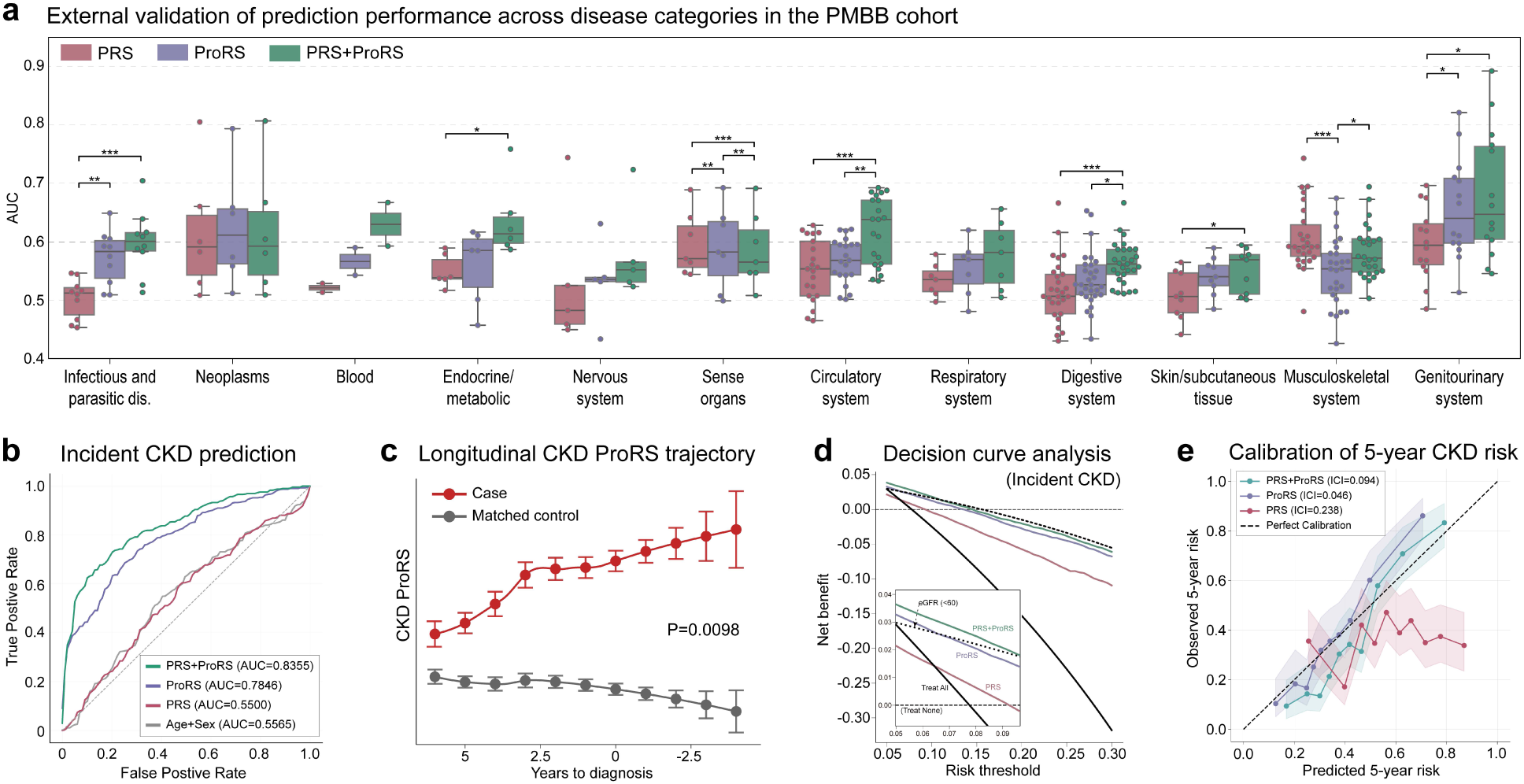
Cross-cohort validation of proteomic and polygenic risk prediction models in PMBB. We evaluated the transferability and clinical relevance of risk models developed in the UKB by applying them to an external cohort with matched genetic and proteomic data. **a**, At the phenome level, prediction performance across 12 disease categories shows consistent improvement when proteomic scores are included. **b–e**, Focusing on CKD as a representative example, we demonstrate how multimodal scores translate into meaningful clinical discrimination (b), temporal risk profiling (c), and decision curve analysis (d), while maintaining strong calibration (e).

To evaluate model performance across ancestral backgrounds, we stratified results by individuals from multiple genetic similarity groups in PMBB, which includes 56.1% African (AFR) and 42.9% European (EUR) ancestry individuals. PRS constructed from EUR-derived summary statistics showed limited predictive value in AFR individuals, consistent with known portability limitations. In contrast, ProRS maintained predictive accuracy across genetic similarity groups, and the combined model consistently outperformed PRS alone. For example, in the AFR subset of the PMBB, ProRS improved prediction for all five cardiometabolic diseases examined, including T2D (AUC increased from 0.516 [PRS only] to 0.617 [combined]), MI (0.607 to 0.671), and cardiomyopathy (0.617 to 0.752) (**Supplementary Fig. 11**). These findings raise the possibility that proteomic scores could help mitigate limitations of polygenic models in globally diverse populations, although further studies are needed to directly assess cross-ancestry performance

## Discussion

Proteomic markers in circulation offer a unique opportunity to capture dynamic biological changes that precede clinical disease onset^1^. In this study, we leveraged large-scale plasma proteomics and genomic data to develop ProRS across 301 disease phenotypes and systematically benchmarked them against PRS. Across 268 traits with at least one informative protein, ProRS outperformed PRS in 88.8% of traits, with consistent gains across cardiovascular, metabolic, and inflammatory diseases. Integration of both modalities further improved prediction, particularly for traits with higher SNP-based heritability. A phenome-wide analysis of cross-trait associations between proteomic and genetic risk scores revealed widespread molecular overlap, with many ProRSs showing significant associations with PRSs for both clinically related and unrelated conditions. Temporal analyses showed that ProRS trajectories increased years before clinical onset, with elevated scores observed more than a decade prior to diagnosis. These findings were robust across follow-up proteomic measurements and replicated in an external cohort, supporting the stability and generalizability of proteomic risk signals.

Notably, one of the most compelling findings was the ability of ProRS to further stratify risk among individuals already in the highest decile of PRS. In traits such as RA, CKD, and AF, ProRS distinguished individuals with substantially higher cumulative incidence from those with lower proteomic risk despite similarly elevated genetic predisposition. This highlights the potential clinical utility of ProRS for identifying subsets of genetically high-risk individuals who may benefit from earlier monitoring or intervention. In addition to risk stratification, ProRS may help guide therapeutic approaches by revealing disease-relevant pathways such as signaling and inflammatory processes that are co-opted early in disease development^35,36^. The complementary resolution offered by ProRS within high-PRS groups underscores its value in layered, precision risk stratification frameworks (**Supplementary Fig. 12**).

While PRSs are widely used to estimate inherited susceptibility to complex diseases, their predictive utility varies across traits and groups defined by genetic similarity, and they cannot capture time-varying risk. Although recent studies have shown the potential of circulating proteins for predicting specific conditions^37–39^, they have largely focused on single diseases and lacked systematic cross-comparison with genetic risk models^40^. Our study addresses this gap by providing a unified, phenome-wide comparison of PRS and ProRS using harmonized pipelines and independent validation. Consistent with prior findings, PRS showed superior performance in neoplastic traits where inherited genetic variation plays a substantial role in disease susceptibility. In contrast, ProRS achieved higher accuracy in diseases affecting circulatory, metabolic, and immune systems, where proteomic changes may reflect early or subclinical pathophysiology. While plasma offers a non-invasive window into systemic biology, it may not capture localized molecular processes occurring in specific tissues. The magnitude of ProRS improvement varied by disease system, with greater gains observed in tissue-specific conditions such as pulmonary or dermatologic disorders. In contrast, more systemic traits—such as cardiometabolic and autoimmune diseases—showed modest gains, potentially due to the dilution of tissue-specific signals in serum-based proteomics. This distinction may inform future efforts to integrate context-specific proteomic or transcriptomic data for improved prediction. Nonetheless, this pattern reinforces the complementary value of genomic and proteomic risk profiling and supports integrated modeling approaches.

A key conceptual advance of this study is demonstrating that ProRS captures dynamic^28^, modifiable components of disease susceptibility not reflected in static germline variation. Unlike PRS, which remains constant over time, ProRS reflects the evolving physiological state and is sensitive to subclinical disease activity^41^. Longitudinal analyses revealed that ProRS levels rose steadily more than a decade before clinical diagnosis in conditions such as CKD, T2D, and RA. This temporal sensitivity underscores the potential of proteomic signatures as early indicators of disease and actionable targets for preclinical risk monitoring. While psoriasis is considered one of the most heritable complex diseases, the relatively lower performance of PRS in this cohort—composed primarily of older adults—may reflect an age-related shift in disease etiology. Specifically, ProRS may be capturing metabolically driven or adiposity-associated subtypes of psoriasis, which are more common in later-onset cases, whereas individuals with high PRS often present with disease earlier in life. However, because our analyses are limited to circulating plasma proteins, ProRS may be less informative for conditions where tissue-specific processes dominate. Integrating tissue-level proteomic data in future studies may enhance risk prediction, particularly in traits where ProRS added limited value. In addition, the analysis of cross-trait associations between ProRS and PRS revealed that individual proteomic risk scores often correlated with genetic risk scores for clinically related or unrelated traits. While these associations reveal correlated risk patterns between proteomic and genetic scores, they should not be interpreted as evidence of biological pleiotropy or causal overlap. Instead, they may highlight domains of converging signals due to shared comorbidities, overlapping feature selection, or systemic disease processes. These findings suggest that proteomic and genetic risk profiles can capture partially overlapping biological processes and provide complementary information for understanding complex disease architecture.

From a clinical perspective, the integration of ProRS and PRS offers opportunities for individualized risk stratification and early detection. ProRS performed best in short-term prediction windows, particularly within 3 to 5 years of baseline, making it well suited for identifying emerging disease risk. Combined models consistently outperformed either modality alone, as illustrated by cumulative incidence gradients across joint PRS–ProRS strata. These findings demonstrate the potential of multimodal models to identify high-risk subgroups with greater precision. Moreover, the reproducibility of selected proteins across time points and cohorts supports the feasibility of building compact biomarker panels for targeted screening, particularly during preclinical stages. Integrating clinical variables alongside PRS and ProRS may further enhance predictive performance by providing context to proteomic signals and help distinguish early disease activity from background physiological variation.

Several limitations should be considered. First, our models were trained using prevalent cases and evaluated on incident cases; this approach assumes that proteomic features distinguishing existing disease also reflect preclinical risk. While this approach is common in risk prediction, it may be less appropriate for episodic or rapidly progressing diseases. Second, our primary analyses were conducted in individuals with high genetic similarity to European reference populations, which may limit generalizability. However, external validation in the PMBB, which includes individuals from multiple genetic similarity groups, supported us to apply the models in a more heterogeneous population. Lastly, follow-up and external proteomic data were limited to reduced protein panels, introducing some heterogeneity, though predictive performance and feature stability remained high.

This study provides a comprehensive assessment of disease incidence risk using both genomic and proteomic data in a large, well-characterized cohort with extended follow-up. By systematically comparing and integrating PRS and ProRS across the phenome, we highlight the complementary nature of inherited and dynamic risk factors and establish a framework for multimodal prediction. Importantly, our analyses were limited to ∼3,000 proteins measurable in circulation, constrained by current coverage of the Olink Explore 3072 panel. Thousands of additional proteins—including those linked to tissue-specific or rare conditions—remain unmeasured. Our results therefore likely underestimate the full predictive potential of proteomics. With the upcoming expansion to ∼5,000 proteins in the UK Biobank and the development of technologies capable of profiling over 10,000 human proteins, future studies will be positioned to further deepen biological insight and predictive accuracy. In parallel, integrating metabolomic data—another dynamic molecular layer now available at scale in UK Biobank—may enhance risk prediction and provide complementary insight into aging, physiology, and early disease processes. Moreover, the application of advanced machine learning methods may enhance predictive performance by capturing non-linear relationships between genetic, proteomic, and clinical variables, potentially improving individual-level risk stratification beyond penalized regression models. At scale, such integrative data and modeling approaches may serve as a foundation for next-generation precision risk prediction and personalized prevention.

In conclusion, ProRS represents a scalable and clinically informative strategy for quantifying dynamic disease risk. When combined with genetic risk scores, ProRS enables multimodal prediction that captures both inherited predisposition and ongoing physiological changes. As proteomic profiling becomes more widespread, these integrative models may support early intervention, targeted screening, and prospective decision-making. Future efforts should prioritize the inclusion of global populations as well as a broader spectrum of diseases, including rare and underrepresented conditions, and explore the utility of these scores in guiding modifiable risk reduction in real-world clinical settings.

## Methods

### Study population and design

The UKB is a large, population-based cohort comprising approximately 500,000 individuals aged 40–69 years at enrollment between 2006 and 2010. Participants provided blood samples and completed detailed baseline assessments including lifestyle, clinical, and anthropometric measurements. Linkage to hospital inpatient, primary care, cancer registry, and death records enabled longitudinal follow-up.

As part of the UK Biobank Pharma Proteomics Project (UKB-PPP), plasma samples from ∼54,000 participants were profiled using the Olink Explore 3072 panel, yielding relative expression levels for 2,923 proteins after initial quality control from UKB. For this study, we restricted analyses to individuals from the randomized proteomics subset who passed proteomic quality control and shared high genetic similarity with European reference populations (EUR), defined by principal component analysis relative to 1000 Genomes reference populations (UKB field ID 22006). After filtering, the final analysis cohort consisted of 39,843 participants, including 21,308 (53.5%) females and 18,533 (46.5%) males, with available proteomic, genetic, and longitudinal phenotype data.

A subset of participants with repeated proteomic measurements from two follow-up visits—the imaging assessment conducted from 2014 onward and the first repeat imaging visit conducted from 2019 onward—was additionally used for sensitivity analyses assessing the temporal stability of risk scores. Proteomic data from these follow-up visits were limited to the initial release of 1,450 proteins, whereas baseline samples included an expanded panel of 2,923 proteins available through subsequent assay batches. These data enabled evaluation of model reproducibility across time points and assay versions. The use of the UK Biobank resource was approved under Application Number 32133.

### Phenotype definition and disease classification

Phenotypes were defined using endpoint definitions aligned with the FinnGen consortium to ensure consistency between the summary statistics used to construct polygenic and proteomic risk scores. Disease diagnoses were ascertained from linked hospital inpatient and health records in UKB using ICD-10 codes (field IDs: 41270 and 41280). The date of blood sample collection (field ID: 53) was used to temporally classify case-control status. Individuals diagnosed with the target disease prior to blood draw were labeled as prevalent cases, while those diagnosed after blood draw were considered incident cases. Controls were defined as participants with no record of the target disease diagnosis throughout the follow-up period. This temporal framework allowed for distinction between baseline disease status and future disease onset, supporting both cross-sectional and prospective prediction analyses. To ensure sufficient statistical power, we included phenotypes with at least 30 incident cases and 30 controls among participants with available proteomic data.

### Plasma proteomics data processing

Circulating protein levels were quantified using the Olink Explore 3072 proximity extension assay, yielding relative expression values for 2,923 proteins after initial quality control performed by the UK Biobank. The initial dataset exhibited ∼17.5% missingness, consistent with expectations for high-throughput proteomic platforms due to technical detection limits and sample quality variation^42,43^. To reduce imputation bias while maximizing data retention, we applied k-nearest neighbor imputation (k=10), which balances local structure preservation and computational efficiency in high-dimensional data. Before imputation, we excluded individuals with more than 54% missing protein values (n=698), based on the inflection point in the distribution of per-sample missingness. We also removed proteins with >30% missingness across participants (n=3) to ensure sufficient information for imputation. After these exclusions, overall missingness was reduced to 9.5%, supporting robust downstream modeling while preserving a large sample size.

### Genotype data and PRS construction

Genome-wide genotype data were available for all participants and imputed using the UKB imputation pipeline. PRS were constructed for 301 disease phenotypes using publicly available GWAS summary statistics from the FinnGen study (release 12; n=500,348). SNP effect sizes were derived using PRS-CS-auto, a Bayesian regression framework with continuous shrinkage priors that accounts for linkage disequilibrium. Variant selection was restricted to ∼1.2 million HapMap3 SNPs. LD reference panels from the 1000 Genomes Project (individuals from European reference populations) were used for PRS-CS-auto. Raw PRS values were computed using PLINK v1.9 as the weighted sum of genotype dosages, with weights provided by PRS-CS-auto. To assess their predictive utility, logistic regression models were fit for each disease phenotype, regressing disease status on PRS while adjusting for age, age², sex, and the first 10 genetic principal components. Trait definitions between FinnGen and UKB were aligned to ensure consistency in phenotype mapping^2^.

### Development of proteomic risk scores (ProRS)

For each disease phenotype, we developed ProRS using data from prevalent cases and a subset of controls at baseline. Incident cases and the remaining controls were held out for model evaluation, enabling prospective assessment of risk prediction in undiagnosed individuals. To ensure balanced and comparable case-control ratios across training and testing, controls were randomly divided in proportion to the number of prevalent versus incident cases. For stability, the training set was constrained to include no fewer than 20% and no more than 80% of the total control pool for any phenotype.

Models were trained using L1-penalized logistic regression (LASSO) as implemented in the glmnet R package^44^. Predictors included age, age², sex, and the first 10 genetic principal components as unpenalized covariates, along with all 2,920 plasma proteins, which were z-score standardized and entered as penalized variables. The regularization parameter was selected via five-fold cross-validation within the training set, choosing the value that minimized cross-validated prediction error. This procedure enabled automatic feature selection while accounting for high dimensionality and multicollinearity among protein markers. LASSO was selected for its ability to perform feature selection in high-dimensional settings, which is particularly relevant for sparse proteomic signals. Alternative regularization methods, such as ridge or elastic net, may be considered in future work.

### Development of combined ProRS+PRS models

To evaluate the additive predictive value of proteomic and genetic risk, we developed combined models incorporating covariates, PRS, and proteomic features. Models were trained using prevalent cases and a subset of controls, and evaluated on incident cases and the remaining controls, consistent with the approach used for standalone ProRS development.

Combined models were fit using LASSO via the glmnet R package. Predictors included age, age², sex, the first 10 genetic principal components, and the disease-specific PRS as unpenalized covariates, along with 2,920 z-score standardized plasma proteins as penalized features. Penalties were applied only to the proteomic predictors, allowing the covariates and PRS to be retained in all models. The regularization parameter was optimized using five-fold cross-validation within the training set, selecting the value that minimized cross-validated prediction error. This framework enabled direct comparison of model performance with and without the inclusion of genetic and proteomic predictors and allowed investigation of their combined contribution to disease risk stratification.

### Model training and evaluation framework

For each disease phenotype, we trained four predictive models: (1) a covariate-only model including age, age², sex, and the first 10 genetic principal components; (2) a PRS-only model including covariates and the disease-specific polygenic risk score; (3) a protein-only model (ProRS) including covariates and all proteomic features; and (4) a combined model including covariates, PRS, and proteomic features. Models (1) and (2) were fit using unpenalized logistic regression. Models (3) and (4) were trained using LASSO to enable variable selection among the high-dimensional proteomic features.

To evaluate generalizability, all models were trained using prevalent cases and a subset of controls and tested on incident cases and held-out controls. To assess whether this training strategy introduced bias, we performed a sensitivity analysis using an alternative approach in which both training and testing used incident cases. Predicted risk scores were highly correlated across the two strategies (median Spearman ρ = 0.71), and the mean change in C-index was negligible, indicating that the use of prevalent cases for model training did not materially affect performance estimates.

### Heritability estimation and contribution analysis

SNP-based heritability (h²) was estimated for each disease phenotype using linkage disequilibrium score regression (LDSC) and applied to GWAS summary statistics from the FinnGen study. Analyses were restricted to ∼1.2 million HapMap3 SNPs to ensure compatibility with PRS construction and reduce biases from poorly imputed variants. LD scores were computed using European ancestry reference panels from the 1000 Genomes Project. LDSC was run with default parameters to obtain trait-level heritability estimates on the liability scale.

To quantify the contribution of polygenic and proteomic risk components to overall model performance, we focused on phenotypes where the combined ProRS+PRS model achieved a Nagelkerke R² > 0.1 in held-out evaluation sets. For each of these phenotypes, we fit Cox proportional hazards models including (1) age, age², sex, and the first 10 genetic principal components; (2) PRS; and (3) ProRS. These models were fit to the same testing data used for performance evaluation (incident cases and controls). We then used sequential analysis of variance (ANOVA) to partition the total explained variance into additive contributions from covariates, PRS, and ProRS. Variance attribution was based on the change in model deviance associated with the inclusion of each component. This analysis enabled comparison of the relative importance of genetic and proteomic predictors across phenotypes, and facilitated stratification of traits by heritability to assess how underlying genetic architecture influenced the added value of proteomic information.

### Cross-phenotype associations between ProRS and PRS

To characterize shared molecular risk across traits, we performed a cross-phenotype association analysis between proteomic and polygenic risk scores. PRS were available for 301 disease phenotypes, of which 268 had corresponding ProRS. We excluded 22 traits based on sex specificity or insufficient ProRS sample size (<15,000), resulting in a final set of 246 traits used for analysis.

For each PRS, we fit linear regression models with each of the 246 ProRS as independent variables, adjusting for age, sex, and the first five genetic principal components. Both PRS and ProRS values were z-score standardized before modeling. Individuals were required to have complete data for the target PRS, candidate ProRS, and all covariates. Because ProRS were constructed using phenotype-specific exclusion criteria derived from FinnGen definitions, the overlapping sample size between PRS–ProRS pairs varied across models. Trait pairs with fewer than 1,000 overlapping individuals were excluded to ensure model stability. Two-sided P-values were calculated for the ProRS term in each model. Associations were retained after multiple testing corrections using the Benjamini–Hochberg procedure to control the false discovery rate (FDR). Analyses were performed independently of disease outcome and were designed to assess molecular risk structure across traits rather than infer causality.

### Risk stratification and cumulative incidence analysis

To evaluate the combined utility of polygenic and proteomic risk scores for individual-level stratification, we conducted cumulative incidence analyses across stratified risk groups for each trait. Analyses were restricted to individuals free of the target disease at baseline, and disease onset was defined using the first recorded diagnosis code after blood collection. We applied three complementary stratification strategies:

(A) Joint binary risk groups: Participants were categorized into four groups based on whether their PRS and ProRS values exceeded the 75th percentile: (1) low PRS and low ProRS, (2) high PRS only, (3) high ProRS only, and (4) high PRS and high ProRS. Kaplan–Meier estimation was used to compute cumulative incidence curves over time for each group. (B) ProRS stratification within high PRS: To assess whether proteomic risk further refines genetic risk prediction, we selected individuals in the top 10% of PRS and divided them into ProRS tertiles. Cumulative incidence was estimated within each subgroup. (C) PRS stratification within high ProRS: A complementary analysis was conducted by selecting individuals in the top 25% of ProRS and dividing them into PRS risk groups, followed by cumulative incidence estimation.

These stratified analyses were performed separately for traits in CKM and AiD domains. Differences in cumulative incidence across groups were used to assess additive and conditional contributions of PRS and ProRS to disease risk trajectories over time.

#### Time-Dependent Prediction and Longitudinal ProRS Trajectories

To evaluate the temporal sensitivity of ProRS, we assessed model performance over varying prediction windows. For each trait, we fit ProRS models to predict disease onset within 3, 5, and 10 years after blood collection, as well as over the full available follow-up period. For each window, incident cases were defined based on diagnosis occurring within the specified time frame, and controls were restricted to individuals without the disease during that interval. Area under the receiver operating characteristic curve (AUC) was used to quantify predictive accuracy for each horizon.

To characterize how ProRS levels change with proximity to diagnosis, we conducted a trajectory analysis using a nested case-control design. For each trait, incident cases were grouped by time-to-diagnosis intervals (e.g., 0–1 years, 1–2 years, …, ≥14 years), and each case was matched to five controls based on sex and age (±2 years). Controls were assigned the same pseudo-index date as their matched case to align observation timing. Within each interval, we computed the mean and standard error of ProRS values and visualized trajectories using locally estimated scatterplot smoothing. To test for monotonic increases in ProRS preceding disease onset, we applied the Mann–Kendall trend test to the difference in mean scores between cases and matched controls across time bins^16^.

#### Sensitivity Analysis Using Follow-Up Proteomic Data

To evaluate the temporal stability of proteomic risk models, we conducted a sensitivity analysis using repeated plasma proteomic measurements from UKB follow-up visits conducted several years after baseline. Proteomic data were available for 1,463 proteins at two later visits: the imaging assessment visit (beginning 2014) and the repeat imaging visit (beginning 2019). A subset of 731 individuals had proteomic measurements available at all three time points (baseline and both follow-up visits).

ProRSs were retrained using the reduced protein panel and redefined training labels based on disease status relative to the follow-up visit date. ProRS models were fit using prevalent cases and evaluated on incident cases within each follow-up window. Model development and testing were repeated independently for each visit, excluding individuals with follow-up proteomic data from baseline training. Analyses were performed for traits with at least 10 incident cases per visit, yielding 45 traits for the first follow-up and 9 for the second. To assess protein-level consistency, we applied principal component analysis (PCA) and UMAP to visualize distributional structure across time points. In addition, we examined longitudinal variation in the 25 most frequently selected proteins from the baseline PRS+ProRS models to evaluate intra-individual temporal dynamics. These analyses allowed us to assess both population-level reproducibility and subject-specific variability in proteomic signatures over multi-year intervals.

#### External Validation in the Penn Medicine Biobank

To assess the generalizability of risk models across cohorts, we performed external validation in 841 participants from the PMBB with available genome-wide genotyping and plasma proteomic data. The median age was 58.7 years, and 47.2% of participants were female. Proteins were measured using the Olink Target 96 Cardiovascular II panel, comprising 92 analytes that overlap with those profiled in UKB.

For harmonized evaluation, ProRS models were retrained in UKB using only the 92 overlapping proteins and applied without recalibration to both the PMBB cohort and a held-out subset of UKB participants with the same reduced protein set. To mitigate potential batch effects, protein measurements were normalized prior to model application. PRSs were recomputed in PMBB using the same FinnGen-derived summary statistics and restricted to a unified set of 1.16 million HapMap3 SNPs.

Phenotyping procedures, covariate adjustments, and statistical models were kept consistent with those used in UKB. Disease incidence was defined relative to the sample collection date, and analyses were restricted to incident cases compared with participants free of the target disease. Genetic ancestry in PMBB was inferred from HapMap SNPs and mapped to 1000 Genomes super-population reference groups. All individuals were retained in the analysis regardless of genetic similarity group.

#### Statistical Analysis and Software

Incident disease prediction was evaluated using both the C-index from Cox proportional hazards models and the AUC. Time-to-event was defined as the duration from blood collection (UKB field ID: 53) to first disease diagnosis, death, or end of follow-up (November 2023), whichever occurred first. For time-dependent ProRS evaluations, we defined case status based on diagnosis within 3, 5, 10 years, or over the full follow-up period. At each horizon, individuals with a diagnosis before the cutoff were treated as cases; those remaining disease-free were considered controls. AUCs were computed for each window to evaluate the waning or stability of predictive performance over time.

To examine longitudinal patterns in proteomic risk, we applied a nested case-control framework. Incident cases were grouped by time-to-diagnosis bins, and each case was matched to five age- and sex-matched controls. ProRS trajectories were visualized using locally estimated scatterplot smoothing (LOESS, span = 0.8), and monotonic trends in group differences were tested using the Mann–Kendall trend test. Interaction between PRS and ProRS was modeled using a multiplicative interaction term in a Cox model; significance was assessed via likelihood ratio test (LRT).

SNP-based heritability was estimated using LDSC, applied to GWAS summary statistics and restricted to HapMap3 SNPs using European reference panels. Correlation between raw PRS and ProRS values was assessed using Spearman correlation without covariate adjustment to evaluate the direct association between genetic and proteomic risk. All statistical analyses and visualizations were performed using R (version 4.4.0) and Python. Cox models and ANOVA were implemented using the survival packages in R, and LDSC was run with default parameters. The full analysis workflow was conducted reproducibly using standardized pipelines and is available upon request.

## Data Availability

All data produced in the present study are available upon reasonable request to the authors.

## Acknowledgements

This work was supported by NIGMS R01 GM138597 and NHLBI R01 HL169458. M.G. Levin was supported by the Doris Duke Foundation (Award 2023-0224) and US Department of Veterans Affairs Biomedical Research and Development Award IK2-BX006551. This publication does not represent the views of the Department of Veterans Affairs or the United States Government. M.G. Levin reports research grants from MyOme and consulting fees from BridgeBio, unrelated to the present work. M.A. Guerraty was supported by Burroughs Wellcome Fund and the Penn Cardiovascular Institute (MG). S.A. Apostolidis was supported by K08AR081929 and a Rheumatology Research Foundation Scientist Development Award.

Use of the UK Biobank Resource in the current study was approved under Application Number [32133]. We thank all the participants and researchers of the UK Biobank and the FinnGen study.

We acknowledge the Penn Medicine BioBank (PMBB) for providing data and thank the patient- participants of Penn Medicine who consented to participate in this research program. We would also like to thank the Penn Medicine BioBank team and Regeneron Genetics Center for providing genetic variant data for analysis. The PMBB is approved under IRB protocol# 813913 and supported by Perelman School of Medicine at University of Pennsylvania, a gift from the Smilow family, and the National Center for Advancing Translational Sciences of the National Institutes of Health under CTSA award number UL1TR001878.

---

**Extended Data Fig. 1.**
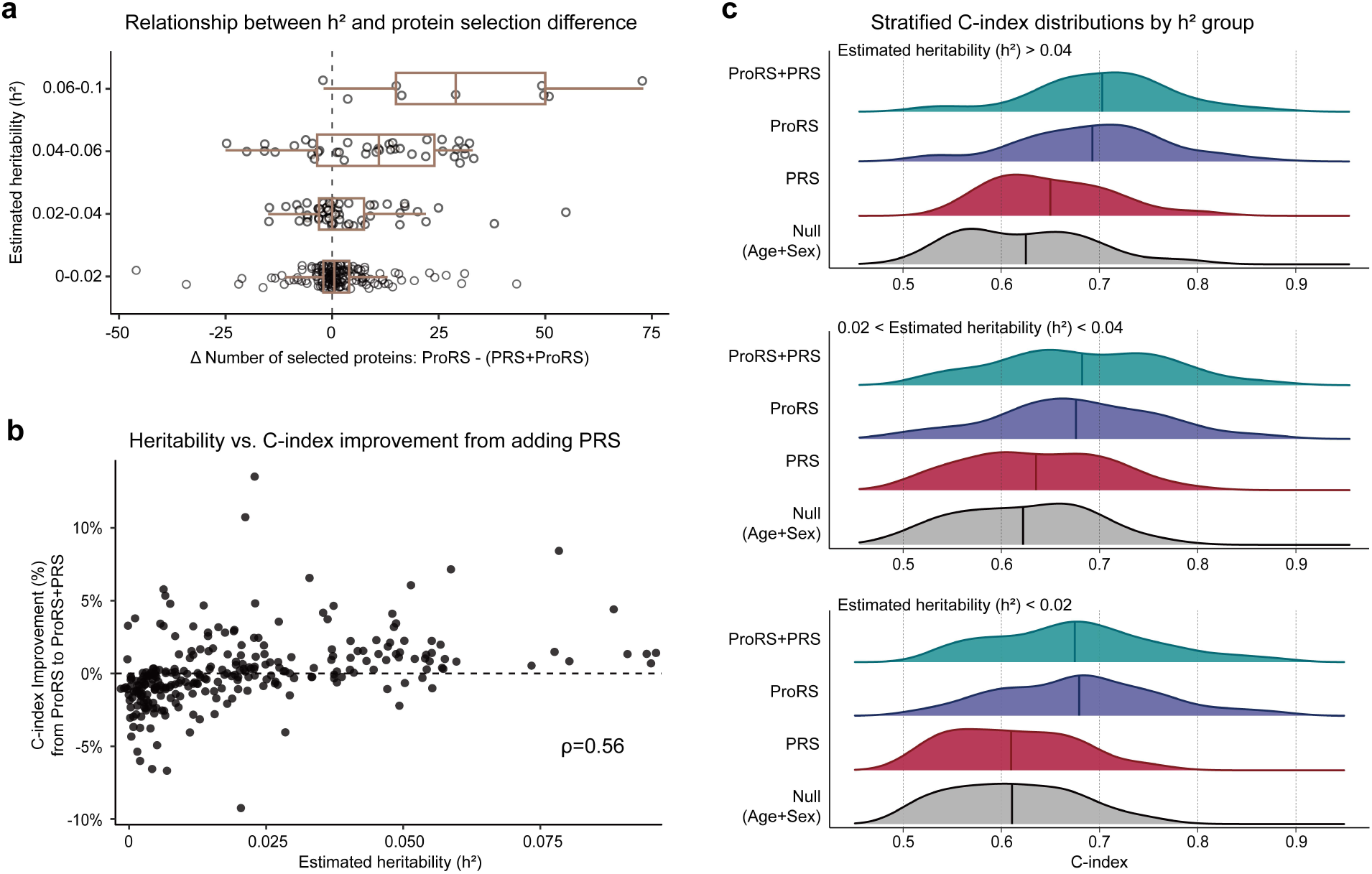
Trait heritability modulates the predictive contribution of polygenic risk scores. **a,** Relationship between SNP-based heritability (h²) and the change in number of selected proteins when adding PRS to the ProRS model. Each point represents a disease phenotype; positive values indicate fewer proteins selected in the combined (PRS+ProRS) model. Box plots summarize protein count change distributions within bins of h², suggesting that PRS partially substitutes for proteomic features in more heritable traits. **b**, Association between heritability and improvement in predictive performance from ProRS to the combined model. Each point represents a phenotype; performance gain was measured by percent increase in C-index. Heritability was positively correlated with C-index improvement (Spearman ρ = 0.56). **c,** C-index distributions for four models (Null: age + sex, PRS, ProRS, and PRS+ProRS), stratified by trait heritability: high (h² > 0.04), intermediate (0.02 < h² < 0.04), and low (h² < 0.02). PRS added substantial predictive value in high-heritability traits but was largely redundant in low-heritability traits.

