## Supplementary Figures for "Large-scale evaluation of proteomic and polygenic risk scores reveals complementary contributions to incident disease prediction"


| Supplementary Figure | Title | Page |
|---|---|---|
| Supplementary Fig. 1 | Phenotype filtering workflow for inclusion in risk score modeling | 2 |
| Supplementary Fig. 2 | Distribution of individual-level Spearman correlations between PRS and ProRS across 268 phenotypes. | 3 |
| Supplementary Fig. 3 | Distribution of protein selection differences between ProRS-only and combined models across phenotypes | 3 |
| Supplementary Fig. 4 | Category-level relationships between heritability and PRS predictive utility. | 4 |
| Supplementary Fig. 5 | Interaction between ProRS and PRS estimates. | 5 |
| Supplementary Fig. 6 | Sensitivity analysis confirms robustness of proteomic predictions at follow-up visits. | 6 |
| Supplementary Fig. 7 | Receiver operating characteristic curve and corresponding AUCs for predicting future diagnosis of cardiometabolic diseases. | 7 |
| Supplementary Fig. 8 | Longitudinal trajectories of the top 25 predictive proteins across UK Biobank visits. | 8 |
| Supplementary Fig. 9 | Histogram of Spearman correlation between ProRS trained from incident cases vs. controls and from prevalent cases vs. controls. | 9 |
| Supplementary Fig. 10 | Histogram of percent improvement in C-index of ProRS trained from incident cases vs. controls and from prevalent cases vs. controls | 9 |
| Supplementary Fig. 11 | Receiver operating characteristic curve and corresponding AUCs for predicting future diagnosis of cardiometabolic disease in the Penn Medicine Biobank | 10 |
| Supplementary Fig. 12 | Joint distributions of PRS and ProRS for six CKM diseases | 11 |

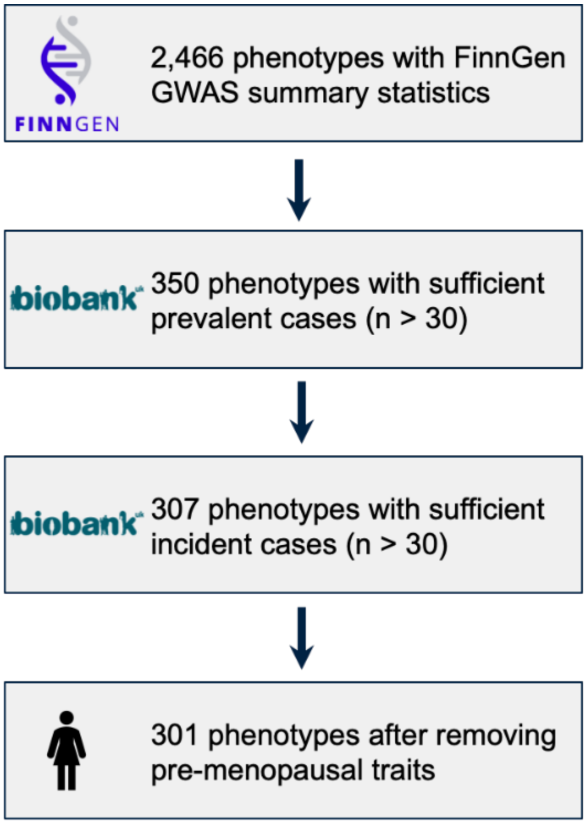


**Supplementary Fig. 1 | Phenotype filtering workflow for inclusion in risk score modeling.** Starting from 2,466 phenotypes with available GWAS summary statistics from FinnGen, we retained 350 phenotypes with ≥30 prevalent cases in UKB-PPP, of which 307 also had ≥30 incident cases. After excluding traits specific to pre-menopausal physiology, 301 binary phenotypes were included for downstream modeling.

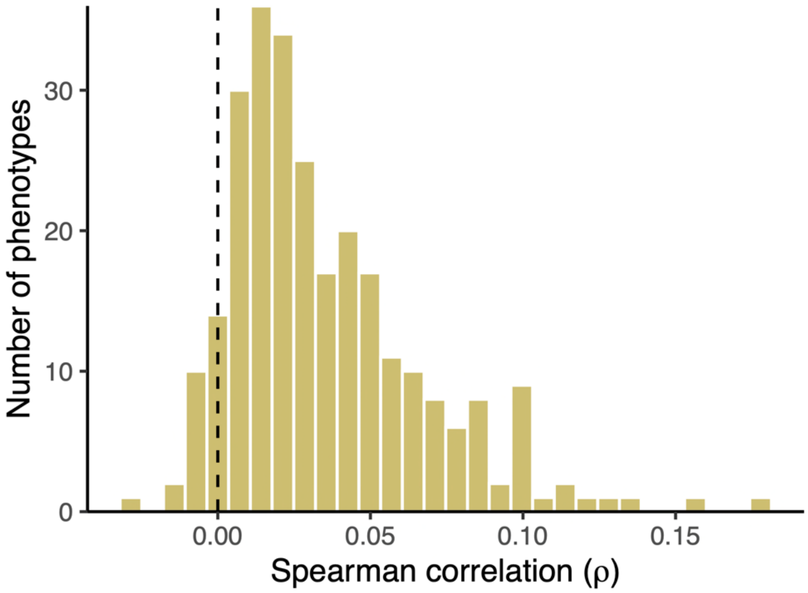


**Supplementary Fig. 2 | Distribution of individual-level Spearman correlations between PRS and ProRS across 268 phenotypes.** Each value represents the within-cohort correlation between PRS and ProRS scores among individuals for a given disease. The majority of correlations are near zero, indicating that the two predictors capture largely distinct sources of variation at the individual level. The dashed line denotes zero correlation.

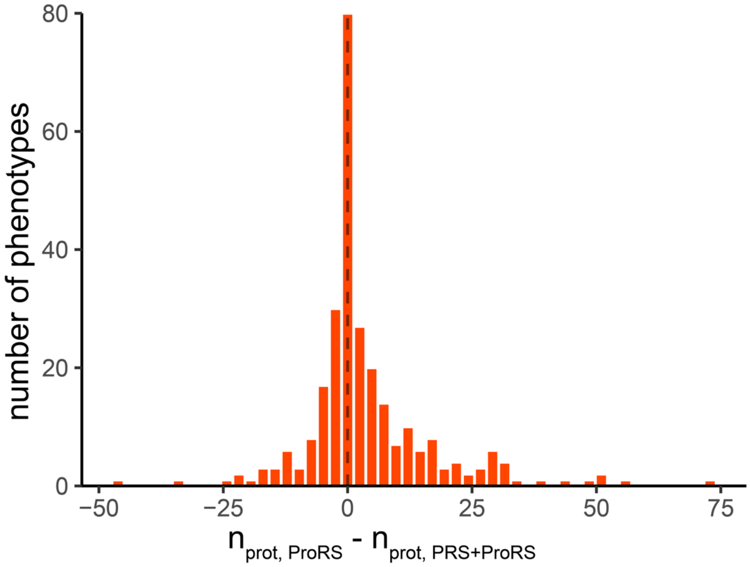


**Supplementary Fig. 3 | Distribution of protein selection differences between ProRS-only and combined models across phenotypes.** Histogram showing the difference in the number of selected proteins between ProRS and PRS+ProRS models across 268 phenotypes (Δ = n_prot, ProRS – n_prot, PRS+ProRS). Each bar reflects the count of phenotypes with a given difference. The dashed vertical line at zero indicates no change in protein count when PRS is added. A modest right skew suggests that genetic information partially substitutes for proteomic features in some traits.

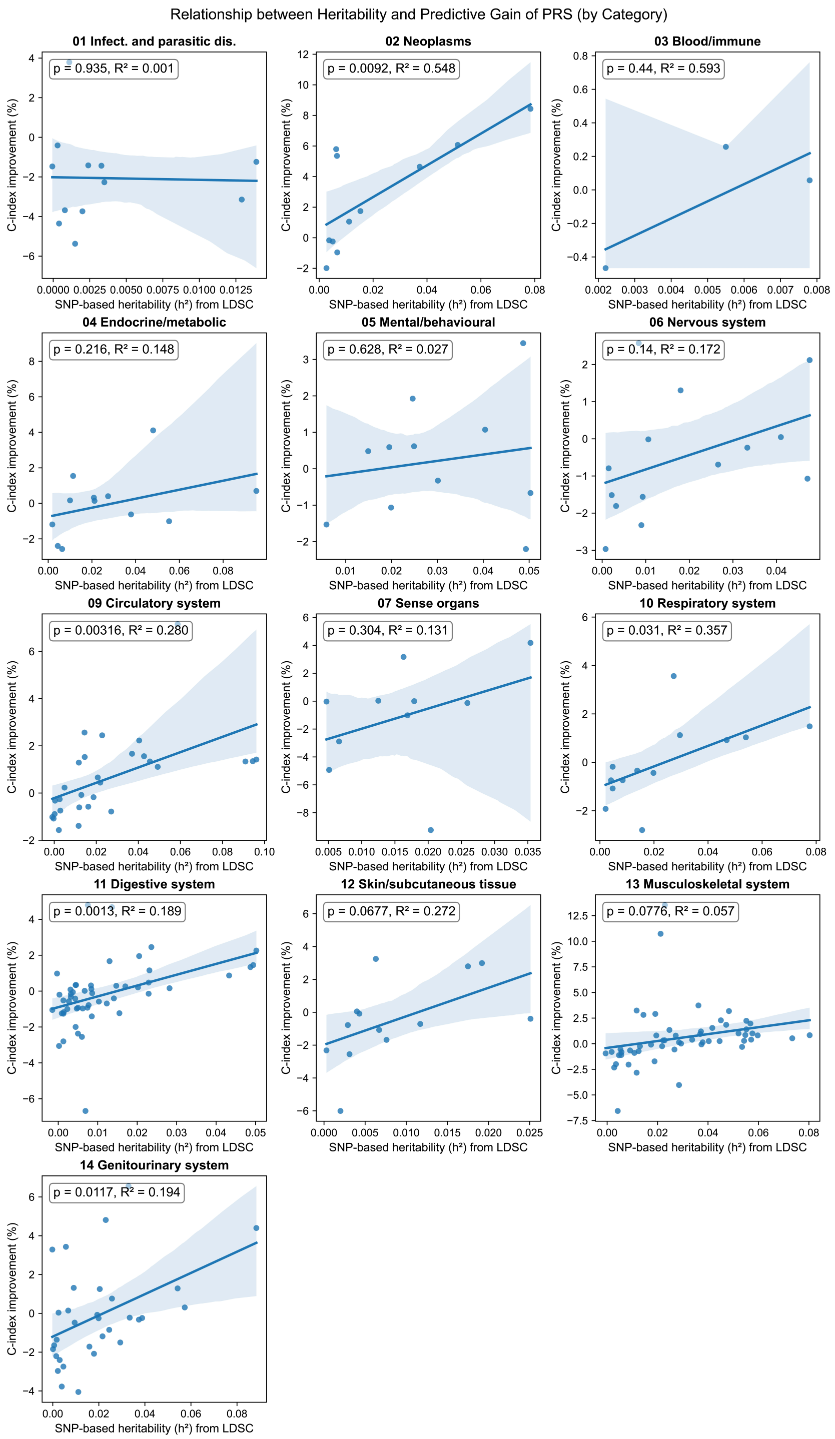


**Supplementary Fig. 4 | Category-level relationships between heritability and PRS predictive utility.** Scatterplots show the relationship between SNP-based heritability ($h^2$; estimated via LD score regression) and the relative predictive improvement from adding PRS to ProRS models (C-index improvement [%]) across 13 clinical phenotype categories. Each point corresponds to a disease phenotype within the category. Trendlines are based on ordinary least squares regression; shaded areas denote 95% confidence intervals. Regression statistics (P-value, $R^2$) are shown per panel. Higher heritability was generally associated with greater predictive gain from PRS addition, particularly in neoplasms, circulatory, and genitourinary disorders.

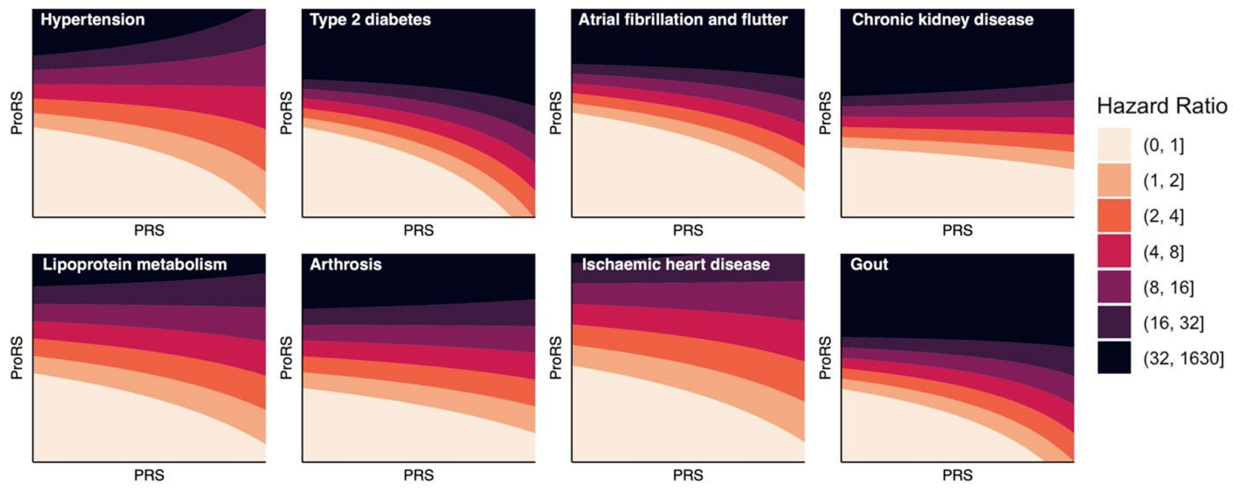


**Supplementary Fig. 5 | Interaction between ProRS and PRS estimates.** Cox proportional hazard models were built using unadjusted PRS and ProRS scores for all diseases, and the hazard ratio change by estimated PRS and ProRS are visualized. Parallel lines indicate less interaction. Phenotypes with the highest interaction, according to likelihood ratio test P-value, are shown: Hypertension (I9_HYPTENS, $P_{LRT} = 1.6 \times 10^{-32}$), Type 2 diabetes (I9_T2D, $P_{LRT} = 1.7 \times 10^{-20}$), atrial fibrillation and flutter (I9_AF, $P_{LRT} = 3.5 \times 10^{-13}$), Chronic kidney disease (N14_CHRONKIDNEYDIS, $P_{LRT} = 2.6 \times 10^{-6}$), lipoprotein metabolism (E4_LIPOPROT, $P_{LRT} = 7.0 \times 10^{-6}$), Arthrosis (M13_ARTHROSIS, $P_{LRT} = 2.3 \times 10^{-5}$), ischaemic heart disease (I9_IHD, $P_{LRT} = 5.8 \times 10^{-5}$), and Gout (M13_GOUT, $P_{LRT} = 1.5 \times 10^{-3}$).

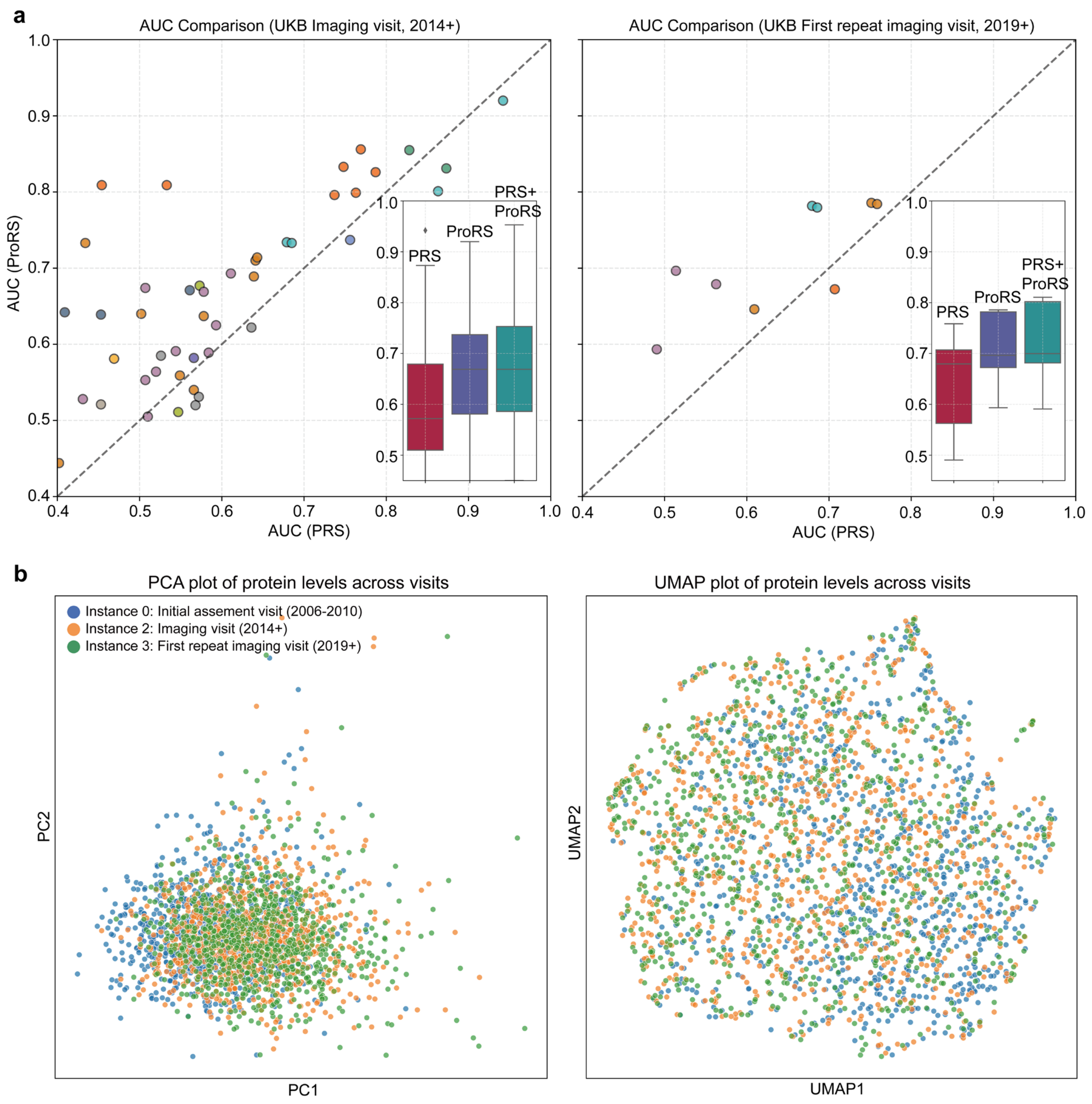


**Supplementary Fig. 6 | Sensitivity analysis confirms robustness of proteomic predictions at follow-up visits. a,** Comparison of AUCs from ProRS, PRS, and combined models at the UK Biobank imaging visit (instance 2, left) and first repeat imaging visit (instance 3, right). Each point represents a disease phenotype; inset boxplots summarize AUC distributions across models. ProRS consistently outperformed PRS despite reduced protein coverage and smaller sample sizes. **b,** PCA (left) and UMAP (right) of 1,450 protein expression values across visits (instances 0, 2, and 3) for overlapping individuals (n = 731). Protein profiles remained stable across time points with minimal batch effects.

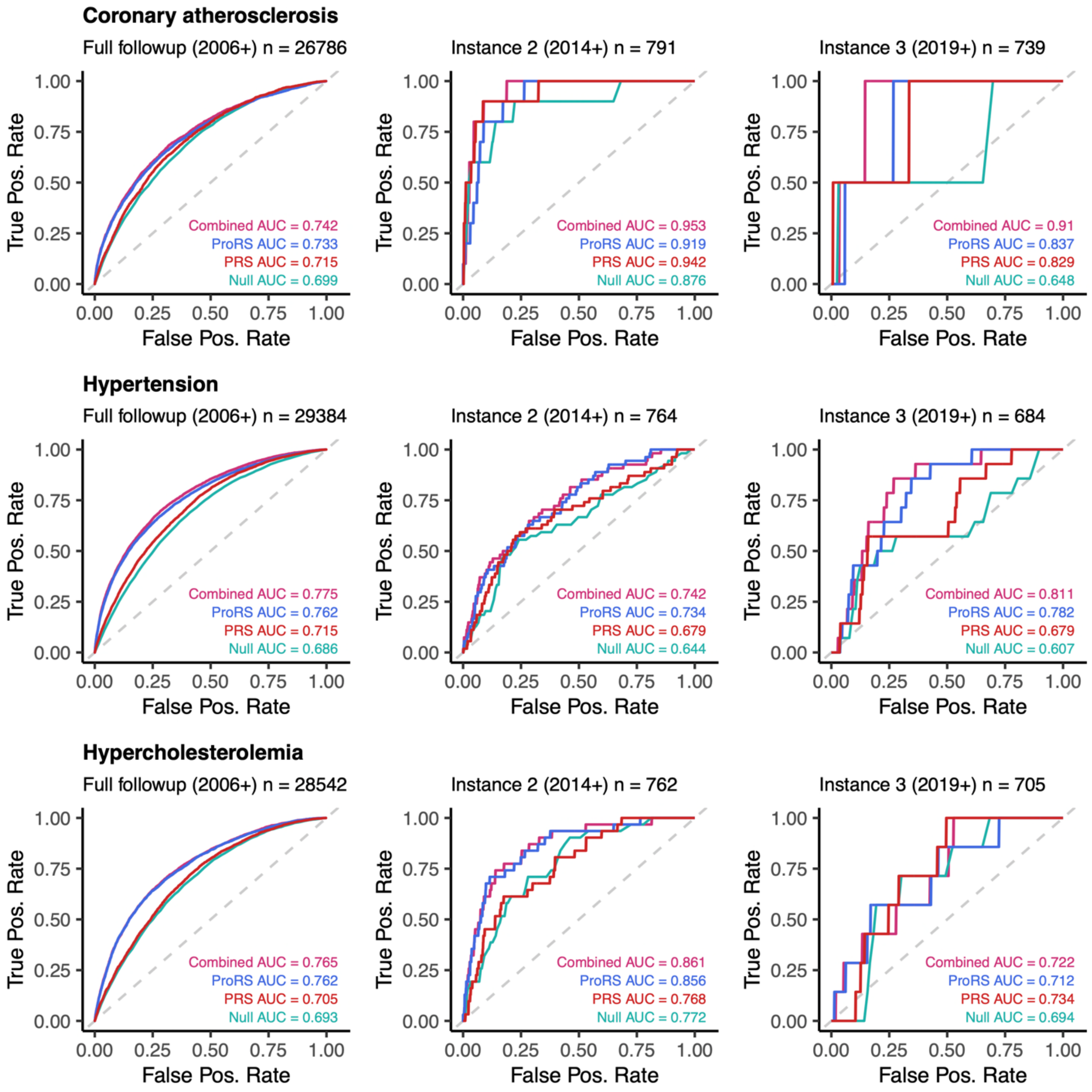


**Supplementary Fig. 7 | Receiver operating characteristic curve and corresponding AUCs for predicting future diagnosis of cardiometabolic diseases** (I9_CORATHER, I9_HYPTENS, and E4_HYPERCHOL) at three different timepoints in the UK Biobank. Individuals with prevalent disease were removed. Combined AUC indicates using both PRS and ProRS, while Null AUC indicates use of just covariates for prediction.

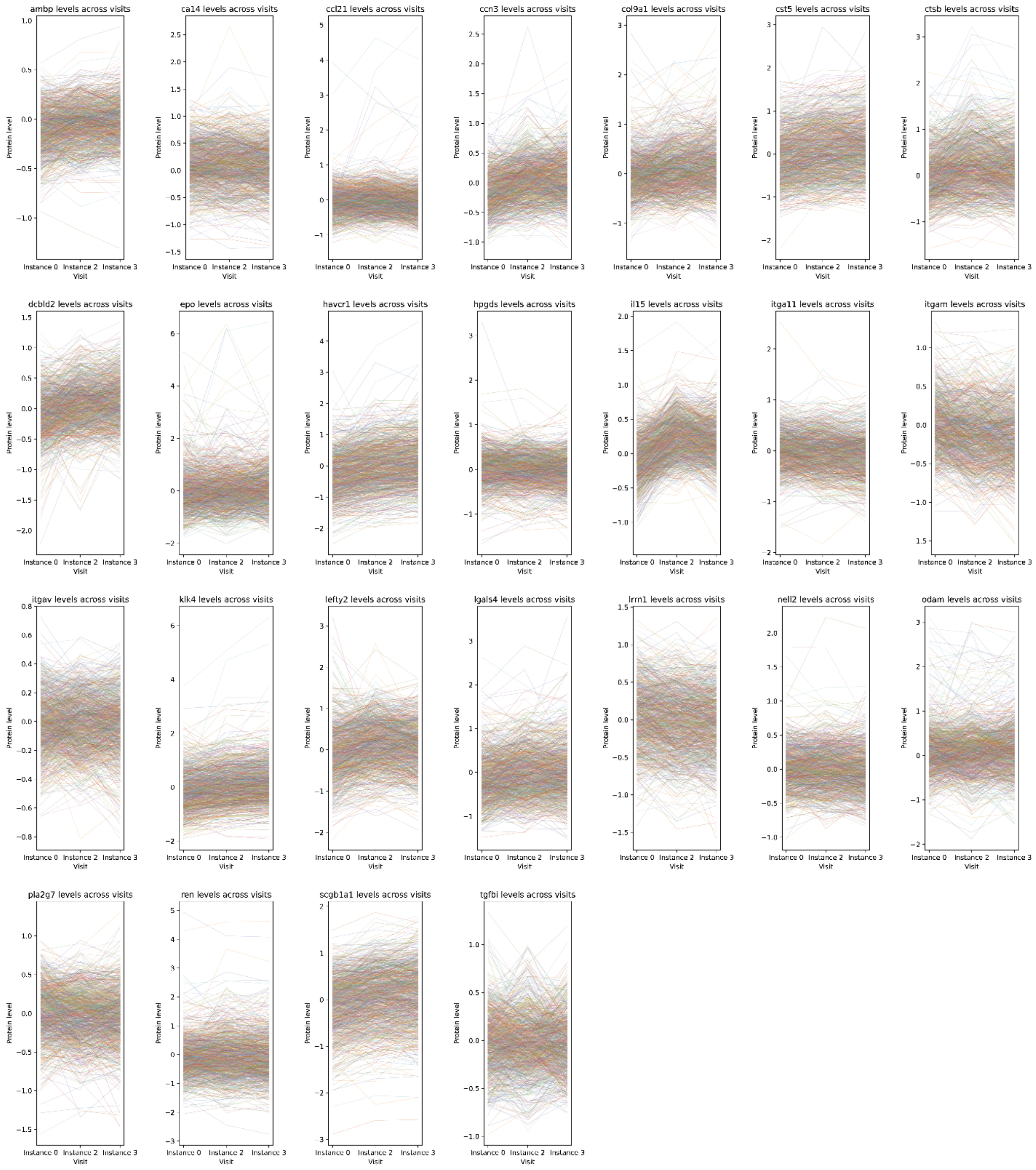


**Supplementary Fig. 8 | Longitudinal trajectories of the top 25 predictive proteins across UK Biobank visits.** Normalized protein expression levels are shown across three time points (instance 0, instance 2, and instance 3) for the 25 proteins most frequently selected in baseline PRS+ProRS models. Each line represents an individual trajectory (n = 731). While population-level expression patterns were stable over time, substantial within-individual variation was observed, reflecting the dynamic nature of circulating proteomic profiles. These findings highlight the temporal sensitivity of proteomics and support its role in capturing evolving health states beyond static genetic risk.

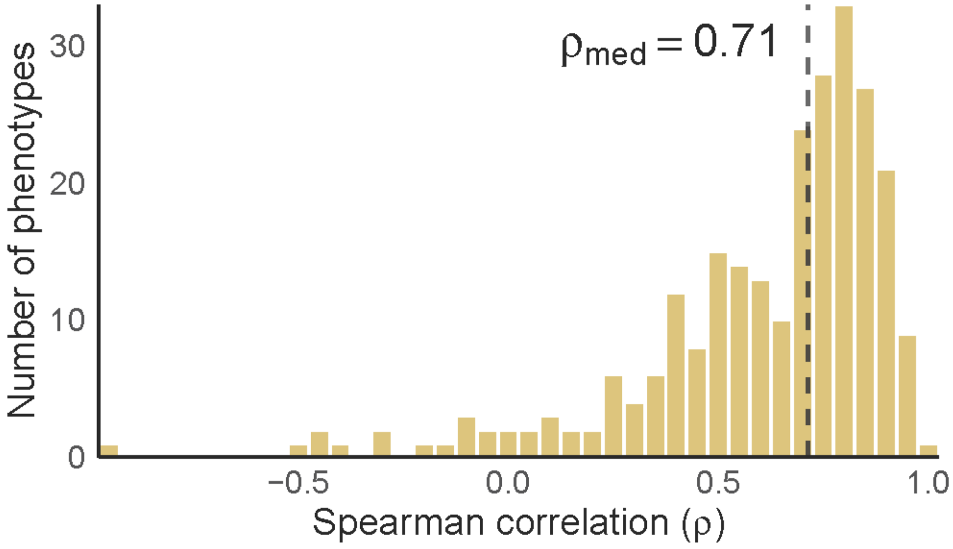


**Supplementary Fig. 9 | Histogram of Spearman correlation between ProRS trained from incident cases vs. controls and from prevalent cases vs. controls.** The mean correlation across all diseases was 0.63.

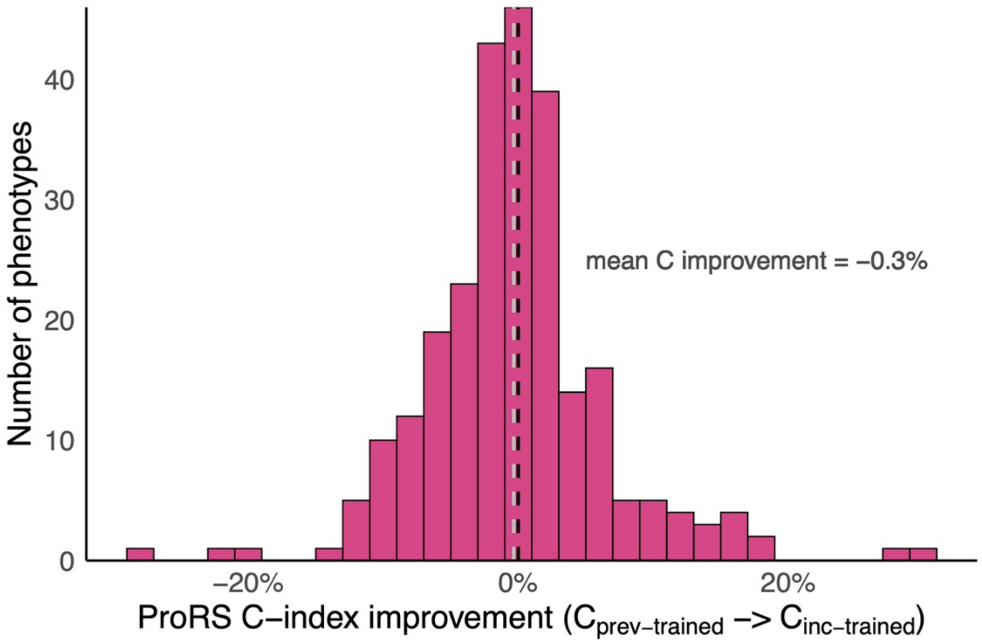


**Supplementary Fig. 10 | Histogram of percent improvement in C-index of ProRS trained from incident cases vs. controls and from prevalent cases vs. controls**. Mean C-index improvement was -0.3%.

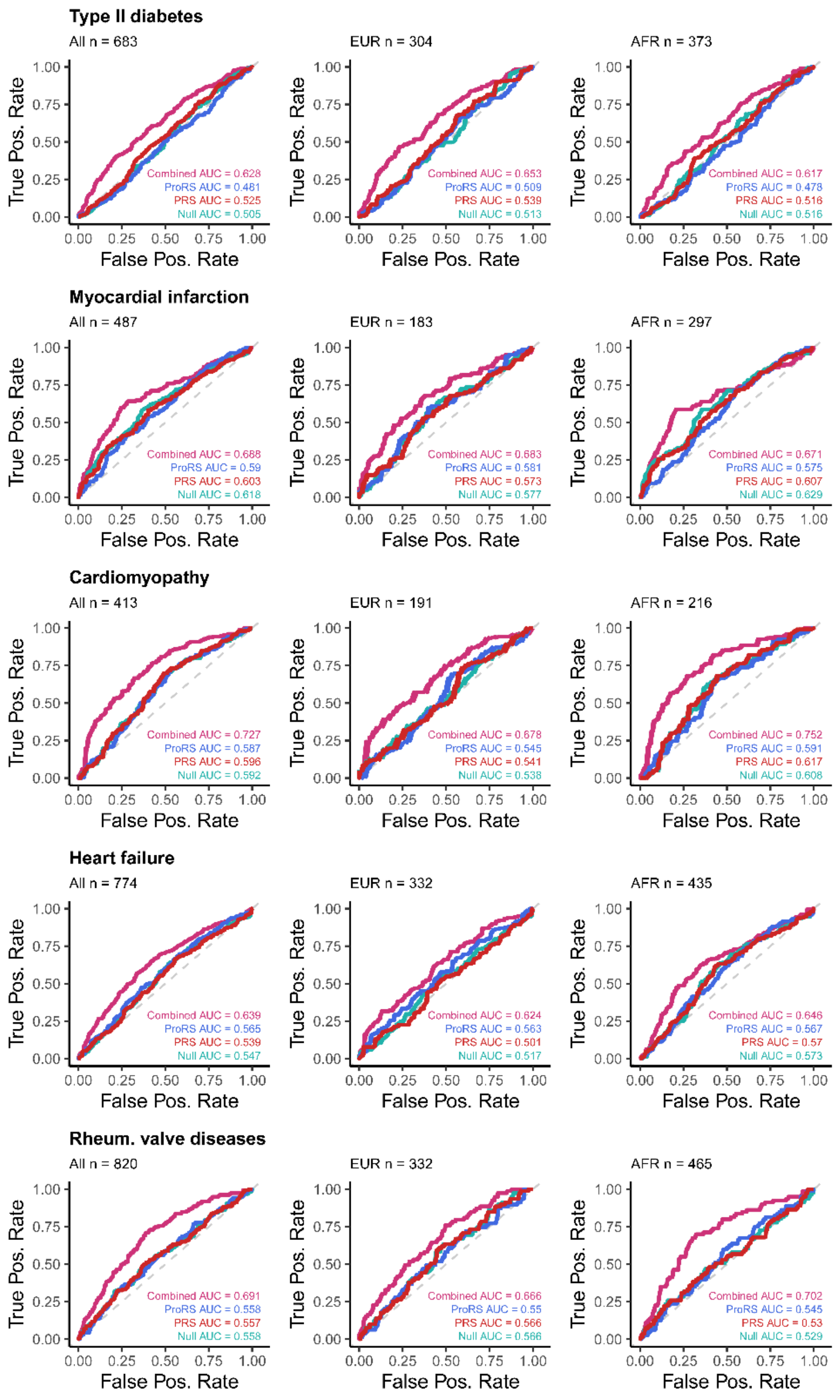


**Supplementary Fig. 11 | Receiver operating characteristic curve and corresponding AUCs for predicting future diagnosis of cardiometabolic disease in the Penn Medicine Biobank,** in the full cohort and just the subset of individuals with genetically predicted European (EUR) and African (AFR) ancestry. Individuals with prevalent disease were removed. Combined AUC indicates using both PRS and ProRS for prediction, while Null AUC indicates use of just covariates for prediction. The traits plotted are type II diabetes (T2D), myocardial infarction (I9_MI_STRICT), cardiomyopathy (I9_CARDMYO), heart failure (I9_HEARTFAIL), and rheumatic valve diseases (I9_RHEUVALV).

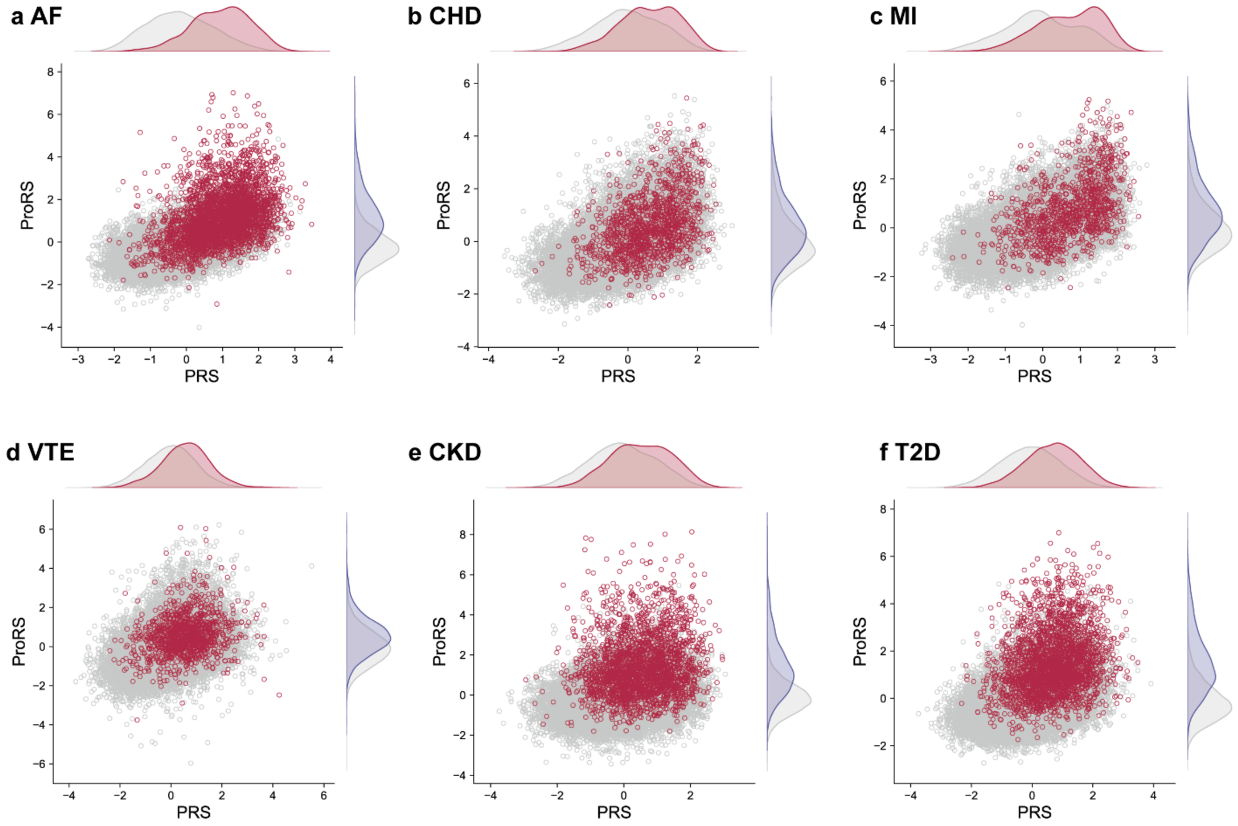


**Supplementary Fig. 12 | Joint distributions of PRS and ProRS for six CKM diseases.** Scatter plots show the relationship between PRS (x-axis) and ProRS (y-axis) in cases (red) and controls (gray) for each trait: (a) Atrial Fibrillation (AF), (b) Coronary Heart Disease (CHD), (c) Myocardial Infarction (MI), (d) Venous Thromboembolism (VTE), (e) Chronic Kidney Disease (CKD), and (f) Type 2 Diabetes (T2D). Marginal density plots indicate the distribution of PRS and ProRS values, highlighting separation between cases and controls.
